# Humoral Responses in the Omicron Era following Three-Dose SARS-CoV-2 Vaccine Series in Kidney Transplant Recipients

**DOI:** 10.1101/2022.06.24.22276144

**Authors:** Caitríona M. McEvoy, Queenie Hu, Kento T. Abe, Kevin Yau, Matthew J. Oliver, Adeera Levin, Anne-Claude Gingras, Michelle A. Hladunewich, Darren A. Yuen

## Abstract

**Background:** Kidney transplant recipients (KTR) have a diminished response to SARS-CoV-2 vaccination in comparison to immunocompetent individuals. Deeper understanding of the antibody response in KTRs following third-dose vaccination would enable identification of those who remain unprotected against Omicron and require additional treatment strategies.

**Methods:** We profiled antibody responses in KTRs pre- and at one and three months post-third-dose SARS-CoV2 mRNA-based vaccine. Anti-spike and anti-RBD IgG levels were determined by ELISA. Neutralization against wild-type, Beta, Delta and Omicron (BA.1) variants was determined using a SARS-CoV-2 spike pseudotyped lentivirus assay.

**Results:** 44 KTRs were analysed at 1 and 3 months (n=26) post-third-dose. At one month, the proportion of participants with a robust antibody response had increased significantly from baseline, but Omicron-specific neutralizing antibodies were detected in just 45% of KTRs. Median binding antibody levels declined at 3 months, but the proportion of KTRs with a robust antibody response was unchanged. 38.5% KTRs maintained Omicron-specific neutralization at 3 months. No clinical variables were significantly associated with detectable Omicron neutralizing antibodies, but anti-RBD titres appeared to identify those with Omicron-specific neutralizing capacity.

**Conclusion:** Over 50% of KTRs lack an Omicron-specific neutralization response 1 month following a third mRNA-vaccine dose. Among responders, binding and neutralizing antibody responses were well preserved at 3 months. Anti-RBD antibody titres may be a useful identifier of patients with detectable Omicron neutralizing antibody response.

**Trial registration:** Clinical Trials Ontario: ID 3604

**Funding:** Funded by the St. Michael’s Hospital Foundation (CMM, DAY) and the Public Health Agency of Canada, through the COVID-19 Immunity Task Force (MAH, MJO, AL).

## Introduction

Solid organ transplant recipients (SOTRs) and patients with chronic kidney disease are at increased risk for severe acute respiratory syndrome coronavirus 2 (SARS-CoV-2) infection and adverse outcomes (1–4). Consequently, kidney transplant recipients (KTRs) represent an especially vulnerable population and a priority group for vaccination. While offering some degree of protection, the humoral response observed following two doses of SARS-CoV-2 vaccine among transplant recipients in general, and KTRs in particular, is inferior to immunocompetent individuals, as is the real world effectiveness of two dose vaccination in this population (5–11).

The improved immunogenicity observed in transplant recipients following a third dose of mRNA vaccine (12–15) led to the recommendation for a primary vaccine series of three doses in this population (16). Subsequent studies have shown augmented binding antibody levels and enhanced neutralizing capabilities following the third vaccine dose in SOTR cohorts, at one and three month timepoints (17, 18). However, diminished responses in KTRs relative to other organ groups have been reported (5, 19), and data on the durability of the antibody response beyond one month in KTR cohorts are lacking.

In late 2021, the Omicron SARS-CoV-2 variant emerged and rapidly gained dominance worldwide. The first Omicron wave related to the BA.1 (B.1.1.159.1) subvariant; however, in recent months another Omicron subvariant, BA.2, has predominated (20, 21). Due to a large number of mutations in the receptor binding domain (RBD) – the main target of neutralizing antibodies – Omicron subvariants can substantially evade the pre-existing humoral neutralization response induced by vaccines or infection (22–26). Importantly, in comparison to a two-dose strategy, a third dose of an mRNA SARS-CoV-2 vaccine has been shown to substantially boost neutralizing antibody titres against Omicron in the general population (25, 27), and to reduce overall infection rates and mortality (28, 29). However, data on neutralizing capacity of KTR plasma to Omicron after additional vaccine doses remain limited. Published studies to-date have focused only on SOTRs (30) or have short (1 month) follow up (31, 32). Moreover, it remains unknown if the correlations previously identified between binding and neutralizing antibodies and protection from infection (33, 34) translate directly to an era in which the highly mutated Omicron subvariants predominate worldwide.

A deeper understanding of the immune response that is predictive of protection from Omicron subvariants would direct future vaccine and pre-exposure prophylaxis strategies in this vulnerable population. Therefore, we conducted a prospective observational study in a cohort of KTRs to comprehensively profile the binding and neutralizing antibody responses at one and three months following the third vaccine dose, and to compare the induced neutralization response with a cohort of healthy controls.

## Results

### Patient characteristics and baseline humoral response

Pre- and post-third dose blood samples were available for 44 KTRs. Baseline demographic and clinical characteristics are shown in **Table 1**. Median age was 55.5 years (interquartile range (IQR), 45.8 to 63 years), and 79.5% of the population was male. The median time from transplant was 43.1 months (IQR, 7 to 142 months). All patients were on a calcineurin inhibitor (CNI), with the majority (91%) taking tacrolimus. 70% were taking an anti-metabolite (mycophenolic acid-based in all cases), 93% of patients were taking prednisone, and in total, 31/44 (70.5%) were on a standard triple-agent immunosuppressive regimen at the time of the third vaccine dose. All participants had previously received two doses of an mRNA-based vaccine, with 35 (79.5%) having received Pfizer-BioNTech BNT162b2, and 8 (18.1%) having been given Moderna mRNA-1273. 26 of the 44 patients (59%) were also tested at 3 months post-third dose. This group had similar clinical and demographic characteristics to the larger parent cohort (**Table 1**).

**Table 1:**
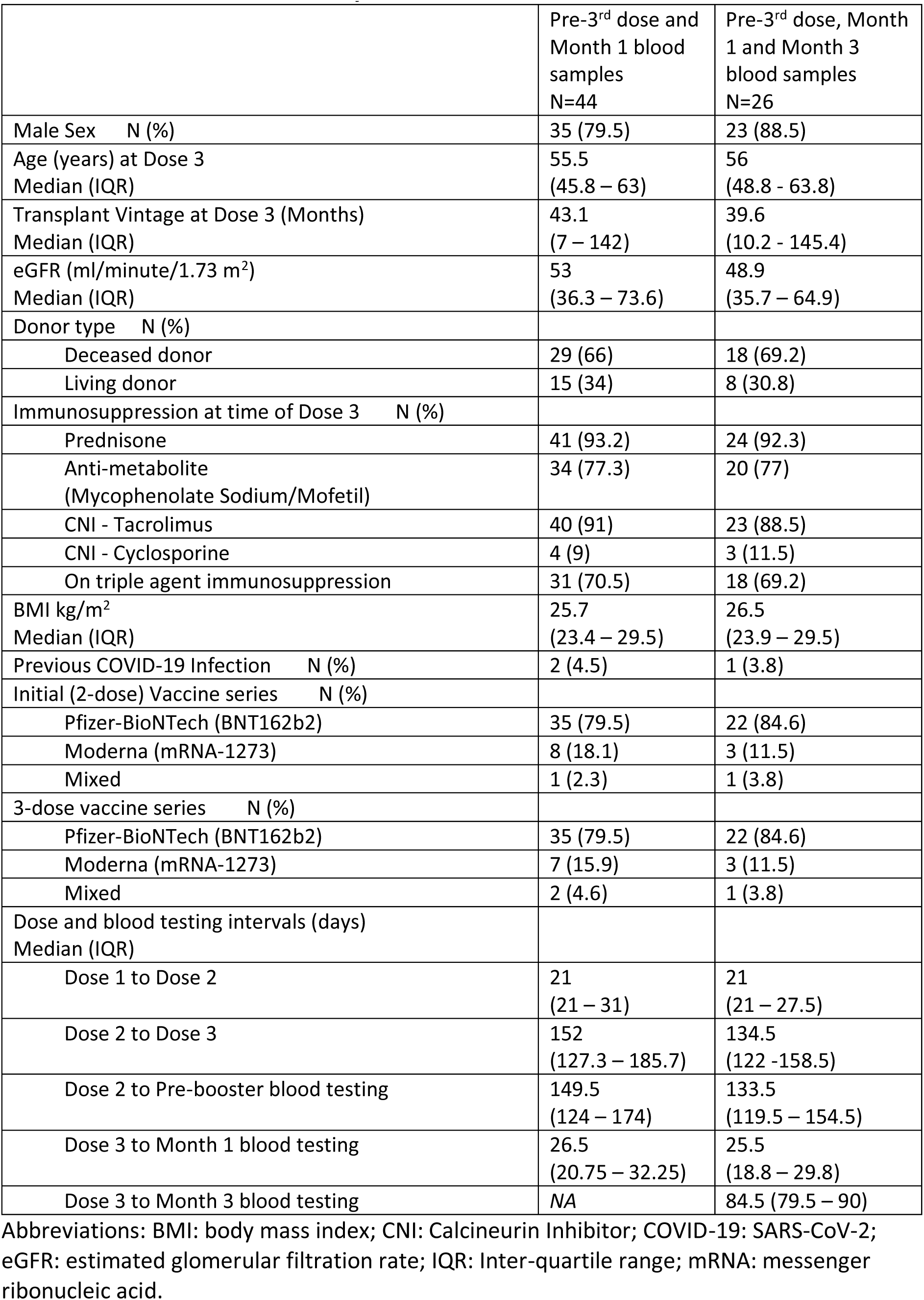
Characteristics of the study cohort.

The baseline demographics of the healthy controls (n= 13) are shown in **Supplemental Table 1**. Their median age was 46 years (interquartile range (IQR), 31 to 55 years), and 30.7% of the population was male. All healthy controls had received the Pfizer-BioNTech BNT162b2 vaccine for their initial two doses.

Anti-RBD, anti-spike and anti-nucleocapsid IgG were measured in plasma before and after a third dose of an mRNA-based SARS-CoV-2 vaccine in KTRs. As described previously (35), the threshold for seropositivity (seroconversion) was determined as ≥3 standard deviations from the log means of aggregated data from archived (pre-SARS-CoV-2 era) negative sera. We considered the median levels of convalescent serum (taken 21–115 days after symptom onset in a cohort of 211 patients in the general population who had mild to severe SARS-CoV-2), as reflective of a robust antibody response (35, 36).

At a median time of 149.5 days (IQR 124-174) post-second vaccine dose, 24/44 (54.5%) and 33/44 (75%) KTRs were seropositive for anti-RBD and anti-spike antibodies, respectively (**Table 2**). However, just 6.8% (3/44) and 18.2% (8/44) of patients had anti-RBD and anti-spike antibody levels consistent with a robust antibody response i.e. exceeding the median convalescent serum levels seen in healthy controls (35) (**Figure 1A-B**). Prior to the third dose, previous SARS-CoV-2 infection had been confirmed by PCR- or rapid antigen-testing in 2/44 patients. However, only 1 of the cases with prior SARS-CoV-2 infection was seropositive for anti-nucleocapsid antibody (**Supplemental Figure 1A**). An additional patient with no known history of SARS-CoV-2 infection was also seropositive for anti-nucleocapsid antibody. In both cases, the titres remained below the median convalescent response observed in healthy controls (35).

**Figure 1:**
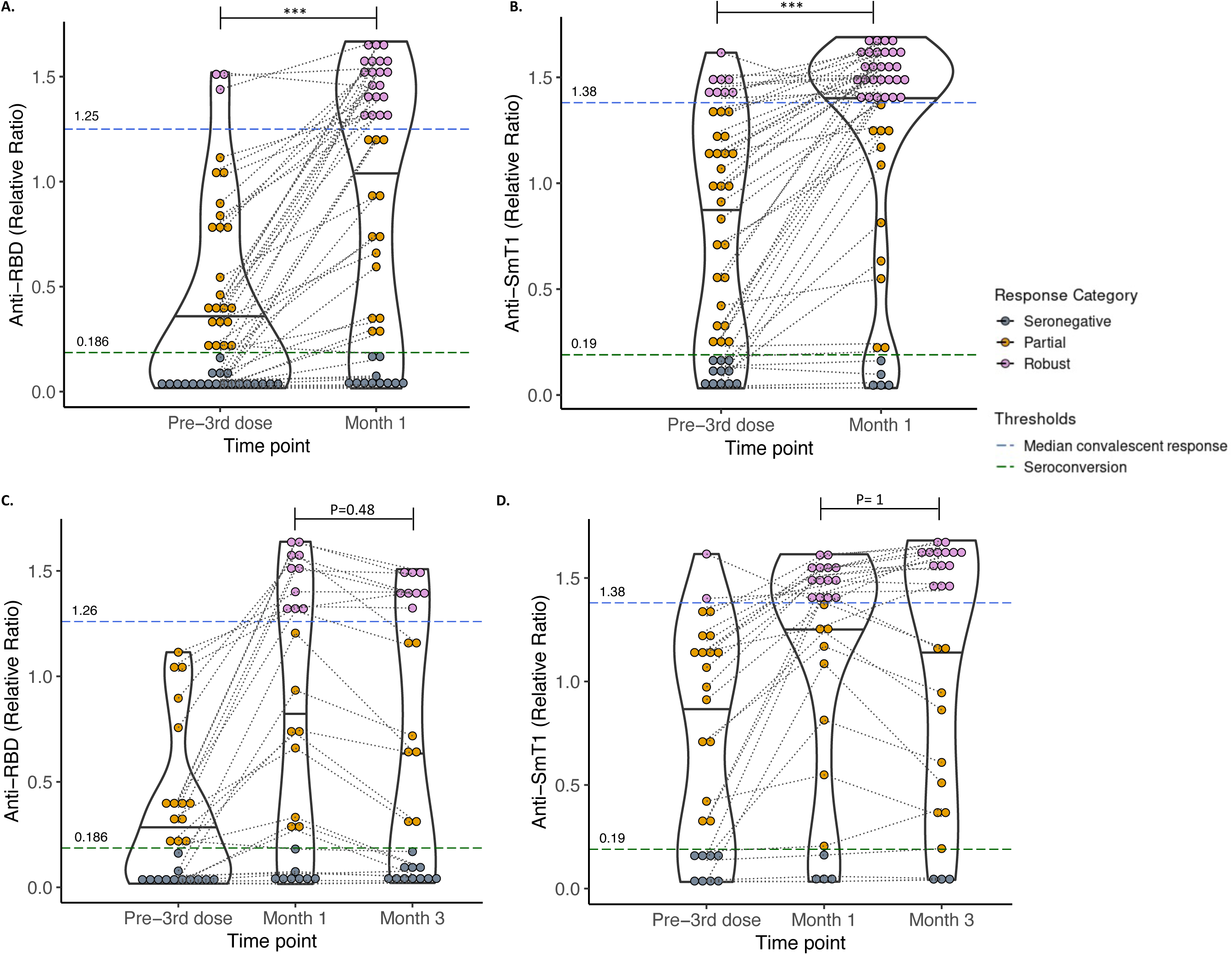
Binding antibody response at 1 and 3 months post-third mRNA vaccine dose. Levels of **A)** serum anti-RBD and **B)** anti-spike IgG in 44 participants with blood samples drawn pre- and at 1 month post-third dose. Anti-RBD and anti-spike antibody levels had significantly increased in the study population at 1 month post-third dose (Wilcoxon signed rank test, p =3.65 x10^-10^ and 7.51 x 10^-10^, respectively). The proportion of kidney transplant recipients now exhibiting a robust antibody response with both anti-spike antibody (McNemar test with continuity correction, p =5.1 x 10-5) and anti-RBD antibody (McNemar test with continuity correction, p =0.001) was significantly increased as shown. Levels of **C)** serum anti-RBD and **D)** anti-spike IgG in 26 participants with additional blood samples drawn at 3 months post-third dose. Anti-RBD and anti-spike antibody levels had decreased at 3 months post-third dose (Wilcoxon signed rank test, p = 8.2 x10^-5^ (anti-RBD) and p = 0.84 (anti-spike)). The proportion of kidney transplant recipients now exhibiting a robust antibody response with both anti-spike antibody (McNemar test with continuity correction, p = 1) and anti-RBD antibody (McNemar test with continuity correction, p = 0.48) was not significantly altered as shown. For all images: The values depicted are relative ratios against a synthetic standard. Serum volume 0.0625 μL. Threshold lines and marked values demonstrate seropositivity (green dashed line) and the median convalescent response (blue dashed line). Individual values are colored to depict the response level as shown in the legend. Solid black lines indicate the median ratio values for each grouping. * p ≤ 0.05, **p ≤ 0.01, ***p ≤ 0.001.

**Table 2.**
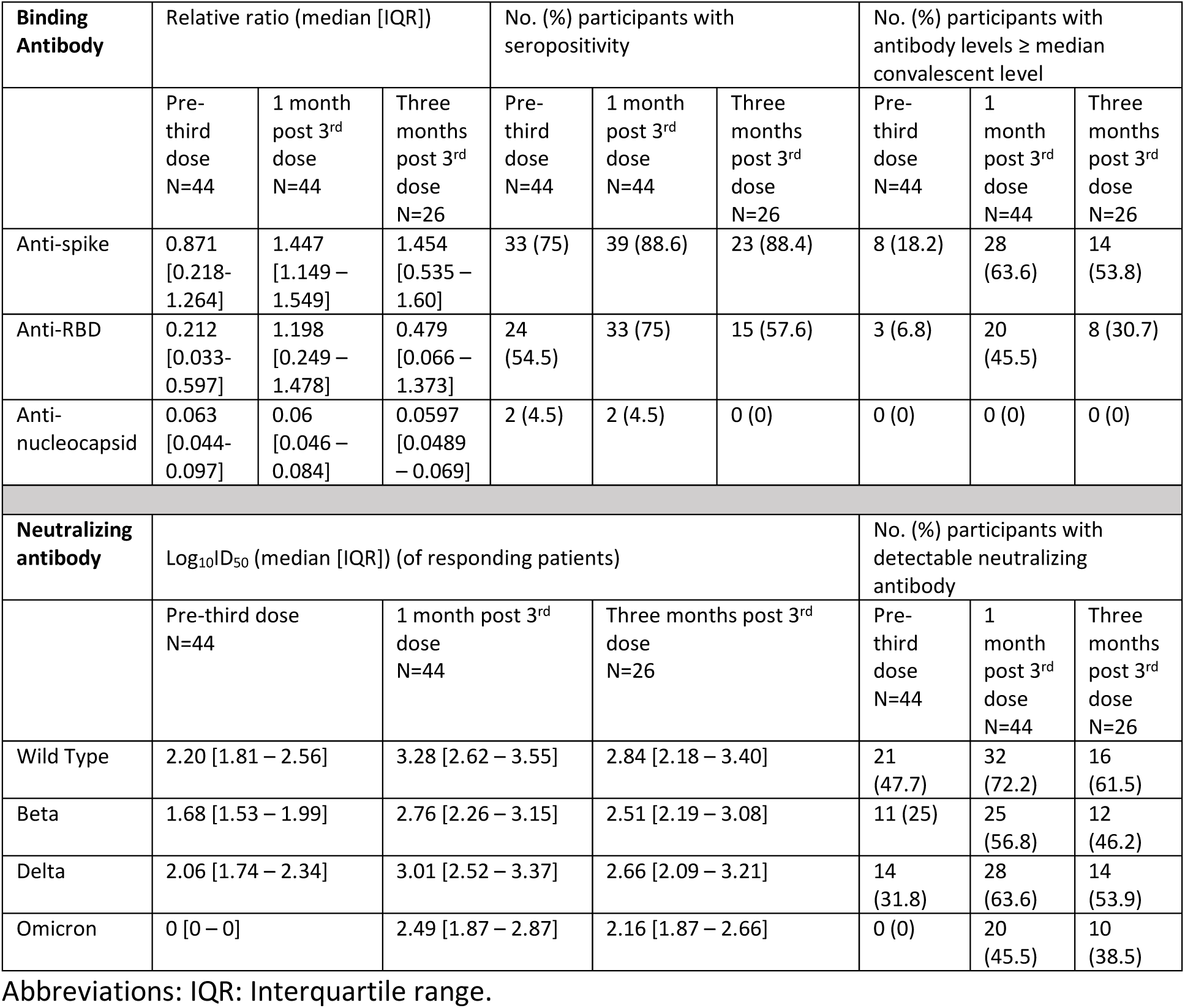
Summary of binding and neutralizing antibody profiles of KTRs prior to, and at 1 and 3 Months after third mRNA SARS-CoV-2 vaccine dose.

### Humoral response at 1 and 3 months post-third mRNA vaccine dose

At a median of 26.5 days post-third vaccine dose, 33 (75%) and 39 (88.6%) KTRs were seropositive for anti-RBD and anti-spike antibodies, respectively. In comparison to pre-third dose serology, anti-spike and anti-RBD antibody levels had significantly increased in our KTR cohort (anti-spike fold increase 1.66, anti-RBD fold increase 5.65; p= 3.65 x10^-10^ and 7.51 x 10^-10^, respectively) (**Figure 1A-B**).

A greater proportion of KTRs now exhibited a robust response, as defined by anti-spike antibody (28/44 (63.6%) participants, p=5.1 x 10^-5^) and anti-RBD antibody (20/44 (45.5%), p=0.001) values above the median levels measured in convalescent healthy controls (**Figure 1A-B**). Although a substantial number of participants remained seronegative at all timepoints tested, 6/11 (54%) and 9/20 (45%) of patients who were seronegative at baseline had seroconverted with respect to anti-spike and anti-RBD antibodies when assessed one month following the third vaccine dose. Anti-spike and anti-RBD antibody levels were highly correlated among individuals (Spearman’s rho 0.82, p < 2.2 x 10^-16^, **Supplemental Figure 1B).**

In total, 35/44 (79.6%) and 7/44 (15.9%) of KTRs received three doses of BNT162b2 (Pfizer-BioNTech) and mRNA-1273 (Moderna), respectively, with 2 KTRs (4.5%) receiving a mixed vaccine series. The majority of those who received mixed vaccine, or Moderna-only regimens had binding antibody titres consistent with a robust response at Month 1; however, differences in proportions were not statistically significant (anti-RBD adjusted p values 0.09 (Pfizer-BioNTech vs. Moderna); 0.14 (Pfizer-BioNTech vs. Mixed); and 1 (Moderna vs. Mixed)), anti-Spike p= 0.59 (**Supplemental Figure 2A-B**). Likewise, anti-RBD and anti-spike antibody levels were not significantly different between Pfizer-BioNTech-only, Moderna-only and mixed vaccination groups (anti-RBD p = 0.1, anti-spike p = 0.22, **Supplemental Figure 2C-D)**.

In 26/44 patients, antibodies were further assessed at 3 months. At a median time of 84.5 days post-third vaccine dose, 15 (57.7%) and 23 (88.5%) of patients were seropositive for anti-RBD and anti-spike antibodies, respectively. There was a significant decrease in the anti-RBD antibody levels of the overall group (p = 8.2 x 10^-5^), while the decline in anti-spike antibody levels was not significant (p = 0.87) (**Figure 1C-D**). Despite the decline in antibody levels, the proportion of the patients in this group exhibiting a robust antibody response with either anti-spike or anti-RBD was not significantly altered (anti-spike: p = 1, anti-RBD: p = 0.48). Exploring this further, we confirmed that participants who mounted a robust antibody response with either anti-spike or anti-RBD antibodies when tested one month post-third dose, experienced a more modest attrition of antibody levels by Month 3 when compared to patients who exhibited a partial response at Month 1, as defined by seropositivity not reaching median convalescent levels in healthy controls (anti-RBD: p =0.012, anti-spike: p = 0.57) (**Supplemental Figure 3A-C**). Similar to the Month 1 response, anti-spike and anti-RBD antibody levels remained highly correlated (Spearman’s rho 0.89, p = 1.6 x 10^-6^, **Supplemental Figure 4A**). Finally, although 1 patient in this group had a prior history of SARS-CoV-2 infection, all patients were seronegative for anti-nucleocapsid antibody at this timepoint (**Supplemental Figure 4B**).

### SARS-CoV-2 Neutralization at 1 and 3 months following third dose

Neutralization capacity at baseline, and at one and three months post-third vaccine dose was assessed for wild-type (WT), B.1.351 (Beta), B.1.617.2 (Delta), and B.1.1.529 (Omicron BA.1) SARS-CoV-2 variants (**Table 2**). Prior to receiving their third dose, 21/44 (47.7%), 11/44 (25%), and 14/44 (31.8%) participants had detectable neutralizing antibodies to WT, Beta, and Delta, respectively (**Figure 2A**). One month post-third dose, the majority of the cohort had detectable neutralizing antibody responses against WT (32/44 (72.7%)), Beta (25/44 (56.8%)) and Delta (28/44 (63.6%)) with median log_10_ID_50_ against WT: 3.28 (IQR 2.62–3.55); Beta 2.76 (IQR 2.26 - 3.15); Delta (3.01 (IQR 2.52 – 3.37)) in the subgroup of positive patients. The proportion of participants with neutralizing antibodies against the WT, Beta and Delta variants had significantly increased compared to baseline (WT: p = 0.026; Beta: p = 0.005; Delta p = 0.005) (**Figure 2A**). No participants had detectable neutralizing antibodies to Omicron prior to third-dose vaccination; however, this was detected in 20/44 (45.5%) of participants after one month (p = 0.0037), with a median log_10_ID_50_ of 2.49 (IQR 1.93–2.86) in the subgroup of positive patients. The proportion of participants with neutralizing antibody against Omicron was significantly lower than those with detectable neutralizing antibody against WT and Delta variants, but not against Beta (p = 0.002 WT versus Omicron, p = 0.074 Beta versus Omicron, p = 0.013 Delta versus Omicron). We assessed the impact of vaccine type on the presence or absence of neutralizing antibodies against each variant; however, no significant associations were identified (WT: p = 0.81, Beta: p = 0.15, Delta p = 0.25, Omicron p = 0.21, **Supplemental Figure 5**).

**Figure 2:**
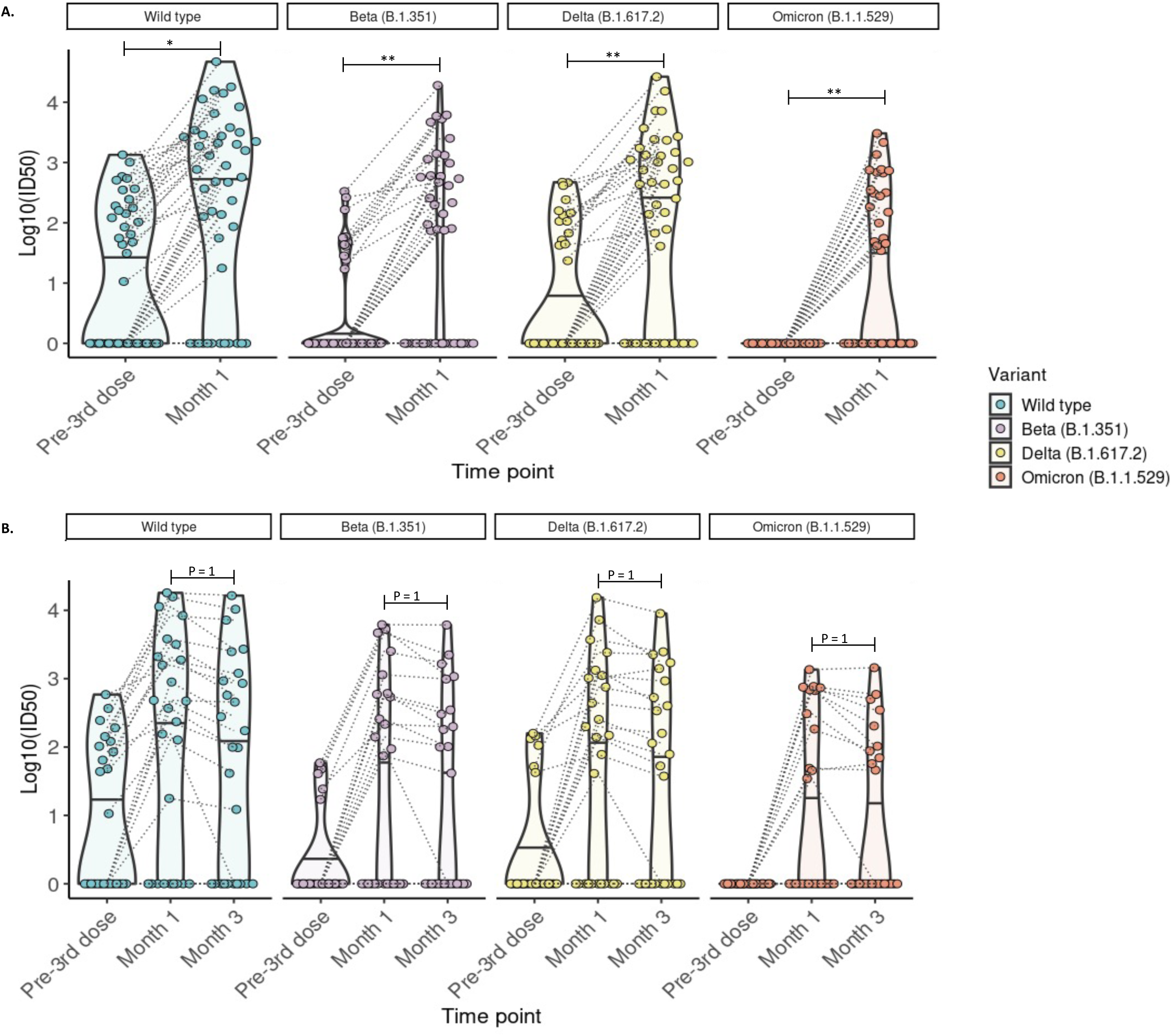
Detection of neutralizing antibodies against SARS-CoV-2 wild-type, Beta, Delta and Omicron (BA.1) variants at 1 and 3 months post-third mRNA vaccine dose. **A)** Neutralizing antibodies detected in 44 participants with blood samples drawn pre- and at 1 month post-third dose. The proportion of kidney transplant recipients with detectable neutralizing antibody (Log_10_ID_50_ >0) was significantly increased for all variants (McNemar test with continuity correction; wild-type p =0.026, Beta p= 0.005, Delta p = 0.005; McNemar’s exact test: Omicron p=0.0037). **B)** Neutralizing antibodies detected in 26 participants with blood samples drawn pre- and at 1 and 3 months post-third dose. The proportion of kidney transplant recipients with detectable neutralizing antibody (Log_10_ID_50_ >0) was not significantly altered compared to month 1 (McNemar test with continuity correction; p=1 for all comparisons. For all images: Paired values are linked with black dashed lines. Solid black lines in each violin plot indicate the median Log_10_ID_50_ values for each variant. * p ≤ 0.05, **p ≤ 0.01, ***p ≤ 0.001.

In the sub-group of patients with blood samples at three months post-third dose, levels of neutralizing antibodies were reduced in comparison to the month 1 timepoint with median log_10_ID_50_ against WT: 2.84 (IQR 2.18–3.40), Beta 2.51 (IQR 2.19 - 3.08), Delta (2.66 (IQR 2.09 – 3.21), and Omicron (2.16 (IQR 1.87-2.66)) in the subgroup of positive patients. Despite this decline, the proportion of patients with detectable neutralization response to individual variants had not significantly altered compared to the month 1 timepoint (p = 1 for all comparisons) (**Figure 2B**). At this time point, a smaller proportion of patients had detectable neutralizing antibodies against Omicron (10/26) as compared to WT (16/26, p = 0.041), Beta (12/26, p = 0.48) and Delta (14/26, p = 0.11).

As compared to WT, neutralizing antibody responses (log_10_ID_50_) were several fold-lower for Beta (median fold reduction 3.4 (IQR 1-7.34), Delta (median fold reduction 1.82 (IQR 1 – 3.50), and most strikingly, Omicron (median fold reduction 11.51 (IQR 1-33.97) in patients with detectable neutralizing antibodies at 1 month and to a lesser extent at 3 months (median fold reductions Beta (2.52 (IQR 1-4.36)), Delta (1.55 (IQR 1-2.62)) and Omicron (6.77 (IQR 1-14.22), **Supplemental Figure 6**).

### Comparison of SARS-CoV-2 antibody responses with healthy controls

The presence and magnitude of binding and neutralizing antibody responses at one month following the third vaccine dose in KTRs was next compared to a cohort of 13 healthy controls. The baseline demographics and vaccination details of this cohort are presented in **Supplemental Table 1**. The magnitude of binding antibody response among healthy controls was increased in comparison to KTRs (anti-spike 1.28 fold increase, anti-RBD 1.58 fold increase). In notable contrast to the KTRs, all healthy controls were categorized as exhibiting a robust response one month after the third vaccine dose (**Supplemental Figure 7 and Supplemental Table 2**).

In contrast to KTRs, neutralizing antibodies against Omicron were detected in 5/13 (38.5%) healthy controls prior to receipt of the third vaccine dose. Additionally, 100% of healthy controls had detectable neutralizing antibodies against all variants, including Omicron, when tested at the 1 month timepoint (**Figure 3** **and Supplemental Table 2**). As observed in the transplant population, neutralization antibody responses were several fold-lower for Beta (median fold reduction 2.26 (IQR 1.62-2.74), Delta (median fold reduction 1.83 (IQR 1.45-2.27), and Omicron (median fold reduction 15.18 (IQR 9.29-18.68), vs. WT at 1 month in healthy controls (**Supplemental Figure 8**).

**Figure 3:**
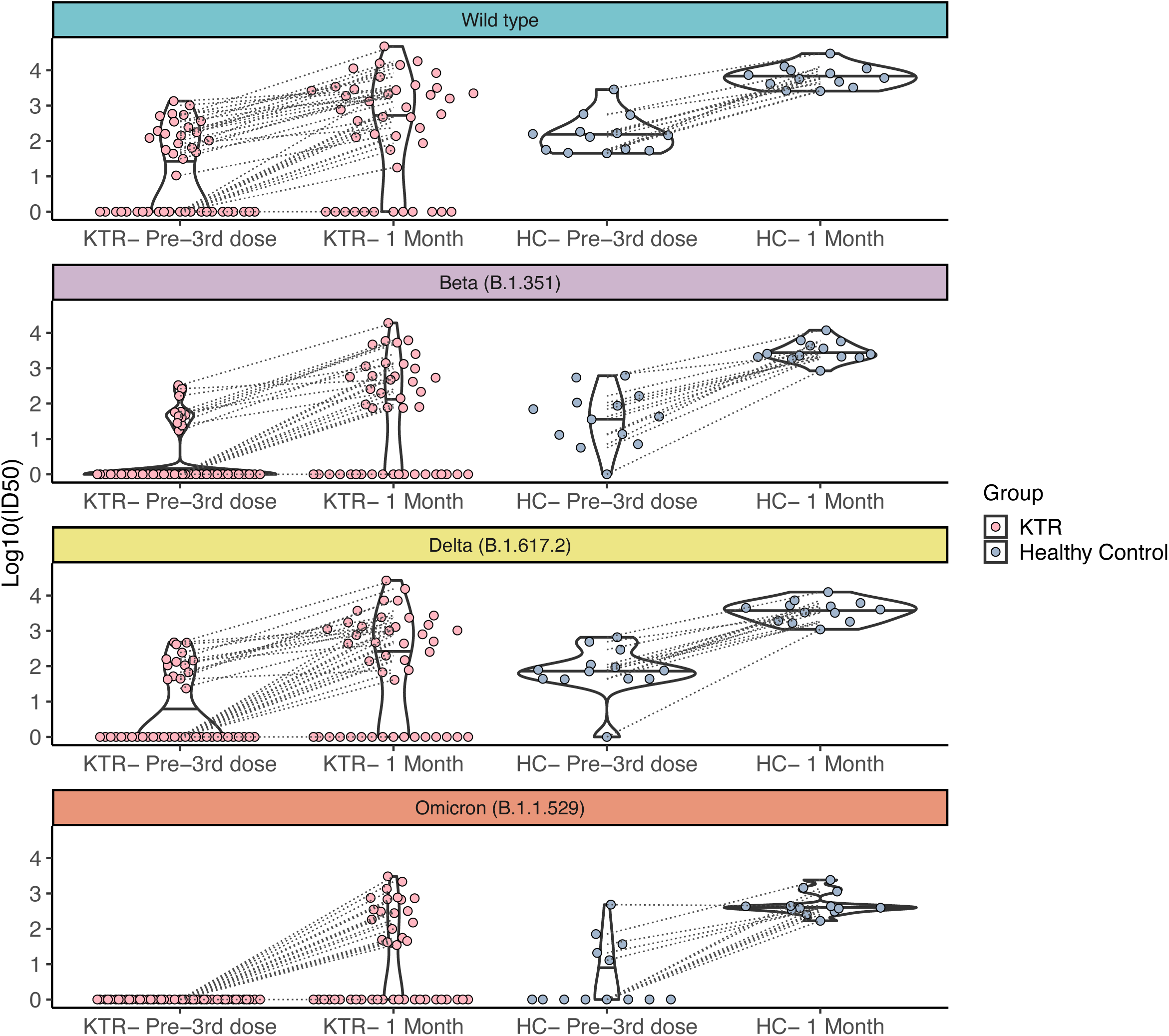
Comparison of neutralizing antibody levels detected in healthy controls and kidney transplant recipients pre- and post-third vaccine dose. Neutralizing antibodies detected in 44 kidney transplant recipients (KTRs) and 13 healthy controls (HC) before the third vaccine dose (pre-third dose) and at month 1 post-third dose. For all images: Paired values are linked with black dashed lines. Solid black lines in each violin plot indicate the median Log_10_ID_50_ values for each variant.

We next focused on the subgroup of ‘responding KTRs’ i.e. those who had detectable neutralizing antibody to an individual variant at 1 month following the third vaccine dose. We compared the median log_10_ID_50_ of healthy controls with that of the responding KTRs, and found that while the magnitude of response in relation to WT, Beta and Delta variants was significantly lower in KTRs (median log_10_ID_50_ fold reduction in KTRs: WT (0.494, p = 0.003), Beta (0.641, p = 0.003), Delta (0.595, p = 0.002)), there was no significant difference in the median log_10_ ID_50_ of responding KTRs and our population of healthy controls against the Omicron variant (log_10_ID_50_ fold reduction in KTRs: Omicron (0.1, p=0.27)) (**Figure 4**).

**Figure 4:**
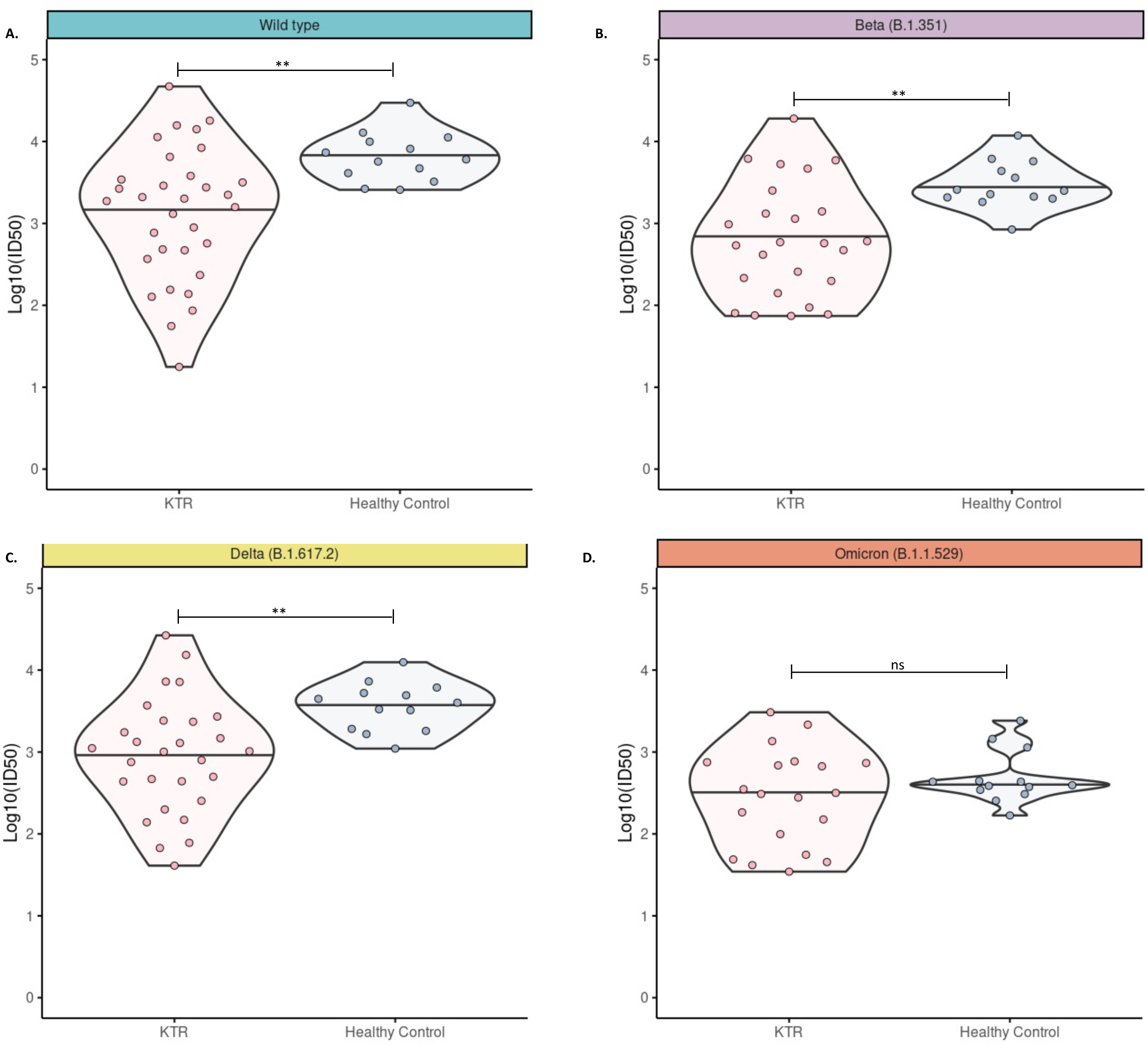
Comparison of neutralizing antibody levels in healthy controls and responding kidney transplant recipients pre- and post-third vaccine dose. For each variant, neutralizing antibody levels for responding kidney transplant recipients (KTRs)- i.e. those with detectable neutralizing antibody (Log_10_ID_50_ >0), were plotted alongside healthy controls (HC) and differences between medians was assessed using Wilcoxon rank sum test. * p ≤ 0.05, **p ≤ 0.01.

### Assessment of responders and non-responders

A number of individuals remained below the threshold of seropositivity for anti-spike and/or anti-RBD antibodies, or had no detectable SARS-CoV-2 neutralization capabilities. As Omicron subvariants are currently dominant worldwide, we reasoned that one of the most important measurable humoral responses would be the presence of Omicron-specific neutralization antibodies. Therefore, we next explored if any demographic and transplant-related clinical factors of our study population associated with the development of Omicron-specific neutralizing antibodies. Individuals lacking a detectable Omicron neutralizing response tended to be older, have a longer transplant vintage, a lower estimated glomerular filtration rate (eGFR), and to have received mainly deceased donor organs; however, no significant associations were identified (**Table 3**). Likewise, no baseline demographic or clinical/transplant characteristics identified people who were non-responders against all variants (n=12), as compared to people with detectable neutralizing antibody against at least one variant (n=32), though responders tended to have a longer interval time between receipt of vaccine doses 2 and 3 (**Table 4**). Serum levels of anti-spike and anti-RBD antibodies were significantly higher in Omicron-specific responders (compared to Omicron non-responders) (anti-spike p = 1.1 x 10^-7^, anti-RBD p = 4 x 10^-7^) and in individuals who had detectable neutralizing antibody against one variant (compared to non-responders for all variants) (anti-spike p = 1.1 x 10^-7^, anti-RBD p = 1.1x 10^-9^) (**Supplemental Figure 9**).

**Table 3:** Characteristics of participant with Omicron-specific neutralizing response.

|  | Non-Responder<br>N= 24 | Responder<br>N=20 | p |
| --- | --- | --- | --- |
| Male Sex N (%) | 19 (79.2) | 16 (80.0) | 1 |
| Age (years) at Dose 3<br>Median (IQR) | 57.50 [51.75, 65.25] | 52.50 [41.25, 58.25] | 0.106 |
| Transplant Vintage at Dose 3 (Months)<br>Median (IQR) | 58.62 [8.12, 131.79] | 24.73 [3.47, 142.16] | 0.316 |
| eGFR (ml/minute/1.73 m<sup>2</sup>)<br>Median (IQR) | 44.30 [36.05, 63.27] | 66.03 [47.92, 79.18] | 0.07 |
| Donor Type (%) |  |  |  |
| Deceased Donor | 19 (79.2) | 10 (50.0) | 0.059 |
| Living Donor | 5 (20.8) | 10 (50.0) |  |
| Immunosuppression at time of Dose 3 N (%) |  |  |  |
| Prednisone | 23 (95.8) | 18 (90.0) | 0.583 |
| Anti-metabolite<br>(Mycophenolate<br>Sodium/Mofetil) | 19 (79.2) | 15 (75.0) | 1 |
| CNI - Tacrolimus | 21 (87.5) | 19 (95.0) | 0.614 |
| CNI - Cyclosporine | 3 (12.5) | 1 (5.0) |  |
| On triple agent immunosuppression | 18 (75.0) | 13 (65.0) | 0.522 |
| BMI (%)<br>(median [IQR]) | 23.91 [22.63, 27.21] | 27.89 [24.77, 31.59] | 0.392 |
| Previous COVID-19 Infection<br>N (%) | 2 (8.3) | 0 (0.0) | 0.493 |
| 3-dose Vaccine series N (%) |  |  |  |
| Pfizer- BioNTech (BNT162b2) | 21 (87.5) | 14 (70.0) | 0.208 |
| Moderna (mRNA-1273) | 3 (12.5) | 4 (20.0) |  |
| Mixed | 0 (0.0) | 2 (10.0) |  |
| Dose and blood testing intervals (days) Median (IQR) |  |  |  |
| Dose 1 to Dose 2 | 21.00 [21.00, 34.75] | 21.00 [21.00, 28.50] | 0.593 |
| Dose 2 to Dose 3 | 140.00 [122.50, 173.75] | 175.00 [130.25, 192.75] | 0.179 |
| Dose 3 to Month 1 blood samples | 25.00 [20.00, 32.25] | 28.00 [21.75, 33.25] | 0.322 |

Abbreviations: BMI: body mass index; CNI: Calcineurin Inhibitor; COVID-19: SARS-CoV-2; eGFR: estimated glomerular filtration rate; IQR: Inter-quartile range; mRNA: messenger ribonucleic acid.

**Table 4:**
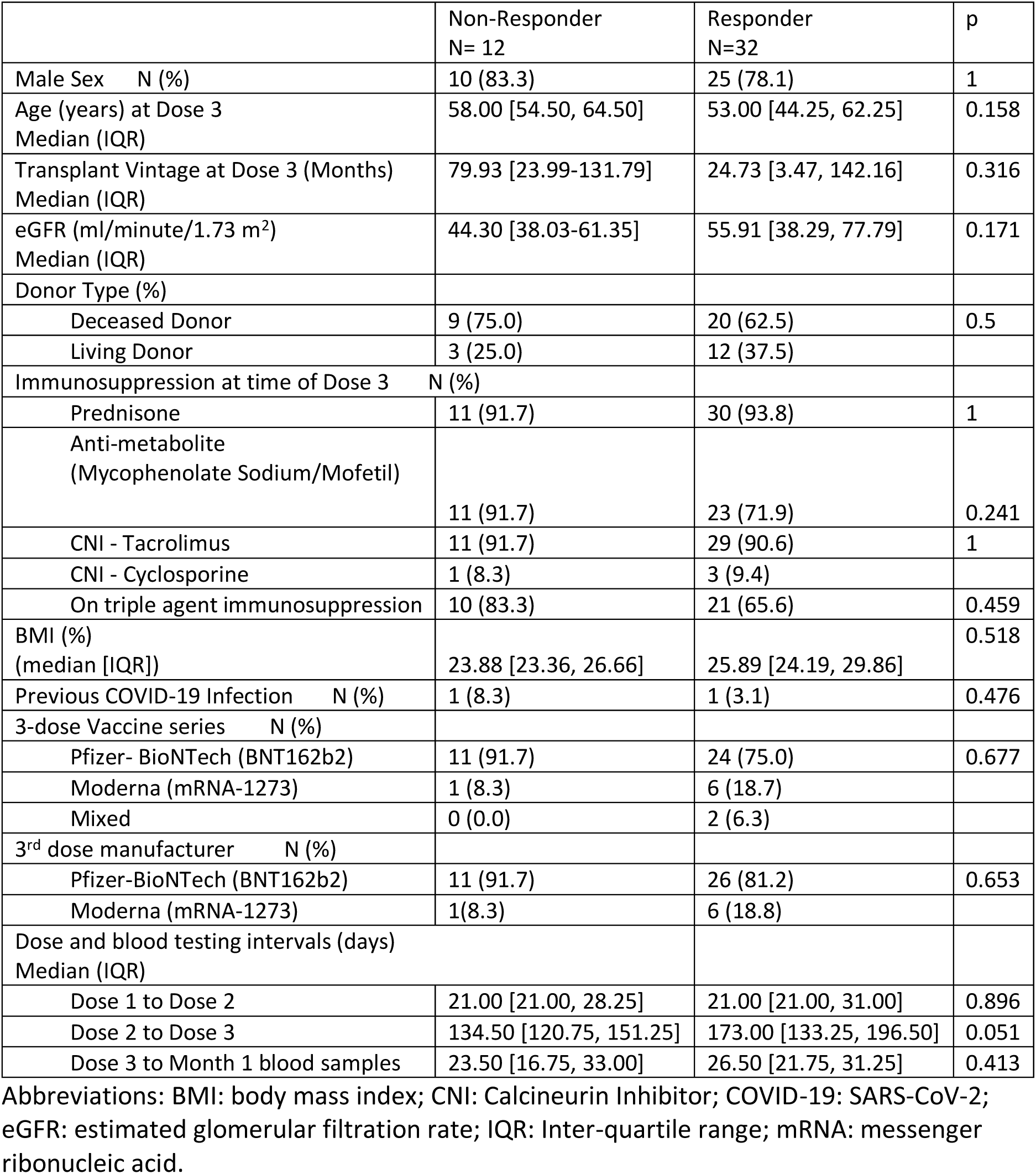
Characteristics of participants with detectable neutralizing response to at least one variant.

As anti-RBD and anti-spike IgG antibody levels at Month 1 strongly correlated with neutralizing antibody levels (Log_10_ID_50_) at both Month 1 and Month 3 (**Supplemental Figure 10**), we next performed receiver operating curve (ROC) analysis to identify the threshold values of anti-RBD and anti-spike antibodies at one month following the third vaccine dose that were associated with the presence of neutralizing antibody against individual variants (**Figure 5**). To maximise specificity, the optimal cut-off was determined by calculating the top-left most point of each curve. For the WT, Beta, and Delta strains, the anti-RBD and anti-spike antibodies had comparable areas under the curve (AUCs), though the thresholds identified for anti-RBD were associated with zero false positives, in contrast to a minimal rate of false positives using the anti-spike threshold (**Supplemental Table 3**). The AUCs were reduced in the case of Omicron (anti-RBD 0.91, anti-spike 0.925), with 3 false positives using the anti-RBD antibody threshold identified, and 4 false positives using the anti-spike antibody threshold identified.

**Figure 5:**
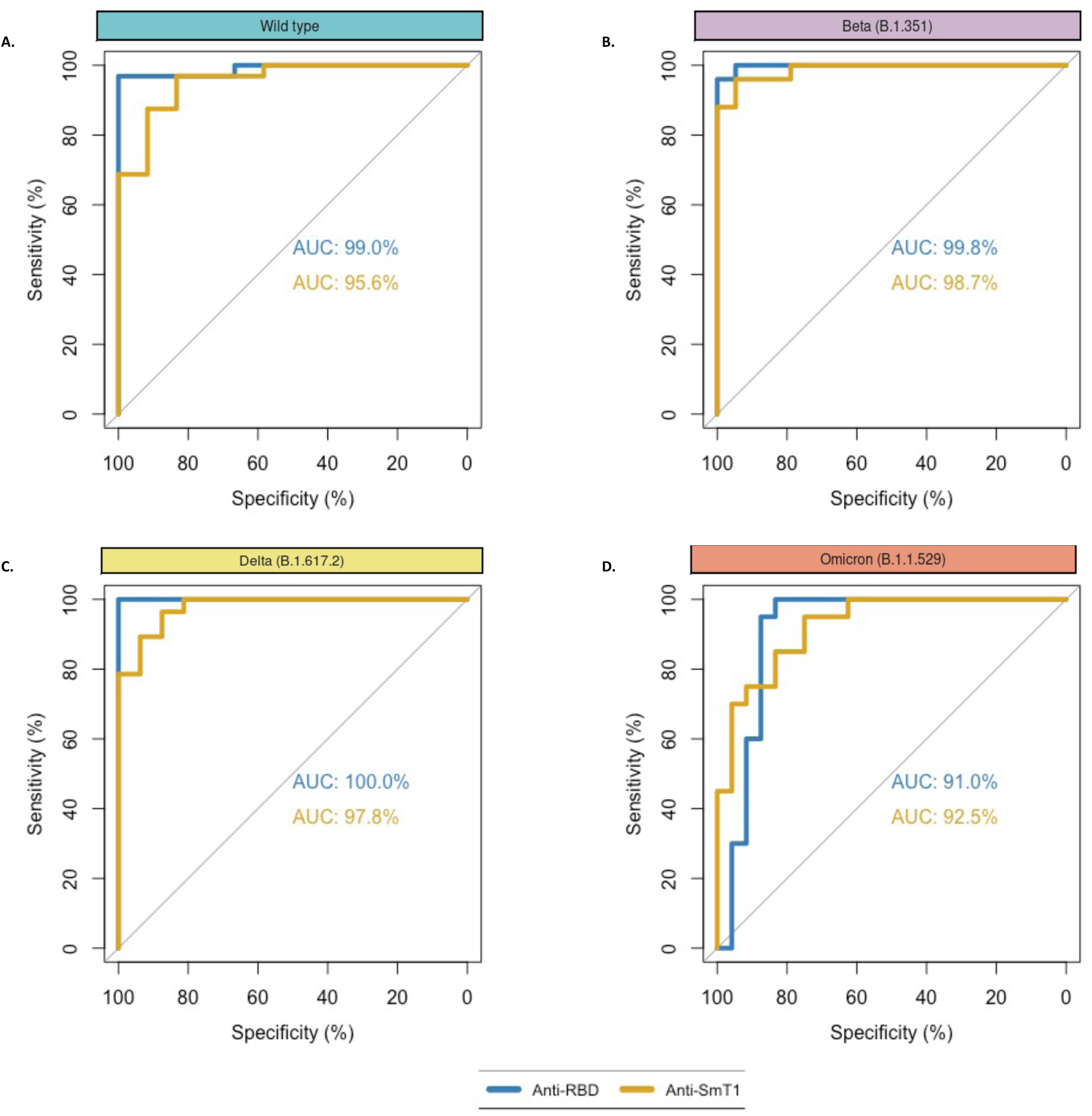
Threshold levels of binding antibody response associated with detectable neutralizing antibody. Receiver operating characteristic (ROC) analysis of anti-RBD and anti-spike antibody levels across **A)** Wild-type, **B)** Beta, **C)** Delta and **D)** Omicron variants, for classification of the presence or absence of detectable neutralizing antibody (Log_10_ID_50_ >0). Areas under the curve (AUC) for anti-spike (yellow) and anti-RBD (blue) are marked. Further details on the threshold values are found in Supplemental Table 3.

The optimal thresholds for anti-spike antibody levels that best identified neutralization capacities against all variants tested (WT: 1.319, Beta: 1.481, Delta: 1.391, and Omicron: 1.475) approximated the median convalescent antibody level seen in recovered healthy controls (1.38). In contrast, the anti-RBD antibody thresholds that best identified neutralization capacity against each variant (WT: 0.317, Delta: 0.627, and Beta: 0.932) were above the seropositivity threshold (0.186), but below the median convalescent response value of 1.25. Notably, in the case of Omicron, the optimal thresholds identified for both anti-spike (1.475) and anti-RBD (1.198) antibodies approximated or exceeded the median convalescent response, suggesting the requirement for a robust level of these antibodies as an identifier of Omicron neutralization capacity.

## Discussion

In this study, we comprehensively profiled the binding and neutralizing antibody responses up to three months following a third dose of mRNA SARS-CoV-2 vaccine in a KTR cohort. The majority of participants had detectable anti-spike and anti-RBD antibodies at one and three months following the third vaccine dose. Our principal findings are: 1) the proportion of patients whose antibody titres were consistent with a robust immune response rose significantly following a booster dose; 2) those who responded robustly at Month 1 had a preserved humoral response at Month 3; and 3) we define anti-RBD and anti-spike antibody levels that may aid in the identification of patients lacking neutralizing antibodies against Omicron, the current dominant variant worldwide.

Our observations regarding overall seropositivity following a third vaccine dose and seroconversion rates in previous non-responders are largely in keeping with the observations in SOTRs (12, 14, 15). Specifically, among previously seronegative patients, 6/11 (54%) and 9/20 (45%) had seropositivity for anti-spike and anti-RBD antibodies when assessed one month post-third dose. While median binding antibody titres had declined at three months, the overall proportion of the cohort who had a robust anti-spike and anti-RBD antibody response was not significantly altered, providing reassuring evidence of a sustained response until this time point (17). Importantly, receipt of a third vaccine yielded a significant increase in the proportion of patients with neutralizing capacity against the WT, Beta, Delta and Omicron variants after one month; however, this response was inferior to that observed in healthy controls. Data on the development of an Omicron-specific neutralizing response among KTRs are limited, with rates of 12% (37) and 43% (38) reported at one month. We detected neutralizing antibody to Omicron in over 45% of our cohort at one month, with sustained response in 38.5% at three months following the third mRNA vaccine dose.

At present, the optimum SARS-CoV-2 vaccination strategy in immunocompromised populations remains unclear. Our findings provide further evidence that the overall humoral response to a three-dose vaccine regimen in KTRs remains inferior to the immunocompetent population, suggesting that alternative strategies, including further vaccine doses, immunosuppressive modulation, or use of complementary agents such as long-acting monoclonal antibodies or anti-virals, may be necessary to induce a protective response against SARS-CoV-2 in transplanted patients (39–41). Concerningly, initial studies suggest that the ‘value-add’ of a fourth vaccine dose may be limited in those with a poor response to a three-dose vaccine series (41, 42). Among SOTRs with a weak serological response to prior vaccine doses, elevated titres of binding and neutralizing antibodies to many variants of concern were noted following a fourth dose; however, neutralization against Omicron was largely unchanged (43). In a KTR cohort, only 10% of previous non-responders after three doses achieved an adequate response following a fourth dose (44). These data suggest that further widespread booster vaccination strategies may prove insufficient to ensure protection from infection, and that an unidentified subset of vaccinated transplant recipients will remain at high risk.

There is a clear imperative to identify correlates of protection from which those with poor serological response could be readily identified. Studies conducted following two-dose vaccination identified a number of clinical factors including age, transplant duration and use of mycophenolate or recent lymphocyte depletion therapies as associated with diminished binding antibody response (45–49). However, in both our study, and the recent study by Kumar et al. in SOTRs (30), no demographic or transplant-related clinical characteristics emerged as significantly associated with the presence of an Omicron-specific neutralization response at one month post-third dose. In our cohort, those with detectable Omicron neutralizing antibody tended to have a lower median age and transplant vintage. In addition, they had a higher eGFR and a greater number of living donor transplant recipients (though these observations are likely linked). Improved vaccine response in proportion to eGFR is well-described amongst individuals with chronic kidney disease (50). Interestingly, patients who had detectable neutralizing antibody against at least one variant tended to have a greater dosing interval between doses 2 and 3, though this did not reach statistical significance. A recent publication by Hall and Ferreira *et al*. found that delayed-interval Pfizer-BioNTech (BNT162b2) mRNA COVID-19 vaccination was associated with enhanced humoral immune responses and robust T cell responses in SOTRs (51).

Both humoral and cell-mediated responses contribute to the development of protective immunity from SARS-CoV-2 (52). The immune correlates of protection are not fully understood; however, binding and neutralizing antibody titres directly correlate with protection from SARS-CoV-2 infection in the general population (33, 34, 53). Functional T-cell responses are also used to infer the degree of immune response to SARS-CoV-2 challenge, yet difficulties with standardization and scalability of these assays have limited their widespread use beyond research settings (54). While neutralizing antibody levels are highly predictive of the extent of immune protection from symptomatic SARS-CoV-2 infection (55), the assays required are typically cell-based, low throughput, and resource intensive in comparison to binding antibody assays (56). Binding antibody assays are higher throughput and more readily scalable, and may thus represent the most accessible means of identifying the subset of patients who remain unprotected (57). Our findings point to binding antibody thresholds which may aid risk-stratification of patients and identification of those requiring prioritization for additional treatment strategies.

As our data and that of others demonstrate, the vaccine-induced Omicron-neutralizing capacity in SOTRs is inferior to that of all other variants (30, 31). The binding and neutralizing antibody thresholds associated with protection from infection represent moving targets, requiring re-definition in the setting of each new variant and associated degree of immune-evasion. A deeper understanding of the duration and magnitude of the immune response in transplant recipients would enable personalized care, and importantly, targeted allocation of scarce resources such as monoclonal antibodies, additional vaccination strategies or novel vaccine compositions within this population.

We acknowledge some limitations to this study. We could not evaluate the associations between neutralizing activity and clinical protection due to a paucity of SARS-CoV-2 breakthrough infections in our population during the study period. Our study size precluded definitive assessment of the impact of mixed vaccine regimens on the magnitude and durability of the antibody response following three-dose vaccination. However, our results, while underpowered, suggest that 3-dose mixed vaccine or Moderna-only regimens elicited stronger humoral responses. Superior immunogenicity from the mRNA-1273 (Moderna) vaccine has been reported amongst both hemodialysis patients and KTRs (36, 58). Further analyses will be required to confirm this result, and to clarify the clinical relevance of this potential finding. Finally, it is likely that the robustness of the protective immune response to SARS-CoV-2 is related to the combined activity of both humoral and cell-mediated immunity, which was not investigated in this study.

In summary, ours is the first study to report on the durability of neutralizing antibodies to Omicron in a cohort of KTRs at three months following the third vaccine dose. Selection of a single anti-RBD-or anti-spike-antibody level threshold whereby neutralizing capability against Omicron is assured is challenging; nevertheless, the majority of KTRs with anti-spike and anti-RBD antibody titres above median levels in convalescent serum had detectable neutralizing antibody, and our work identifies specific binding antibody thresholds which may aid in the risk-stratification of patients. Importantly, quantitative binding antibody assays are relatively high throughput and scalable, making them a potentially attractive option worth further investigation (35).

Whilst future studies will be required to explore the contribution of cell-mediated immunity, affinity maturation and broadly neutralizing antibodies in this setting, our data suggest that antibody levels may aid in the identification of KTRs who remain highly susceptible to infection after vaccination, and for whom additional therapies may be necessary.

## Methods

### Patient population and study design

We conducted a prospective observational cohort study of kidney transplant recipients followed by the Renal Transplant Program of St. Michael’s Hospital (Unity Health Toronto), Toronto, Ontario. All adult patients (aged ≥ 18 years) who had received a kidney transplant and had a functioning allograft were considered eligible for inclusion. Prevalent and incident patients were contacted in September 2021 when eligibility for third vaccine dosing in solid-organ transplant patients was confirmed by the Ontario Ministry of Health, and invited to participate in the study.

### Clinical data sources

Patient demographics, clinical characteristics and covariates, and data related to COVID-19 vaccination or infection status were collected from the St. Michael’s Hospital electronic health records.

### Healthy controls

Healthy controls included health care workers and research staff recruited at Sunnybrook Health Sciences Centre (CTO No. 3604). All healthy controls were in self-reported good health and were ≥ 18 years old. Individuals with chronic conditions including chronic kidney disease, diabetes mellitus, hypertension, HIV, usage of immunosuppressive medication, and prior organ transplantation were excluded as healthy controls.

### Serologic assays

Anti-spike, anti-receptor binding domain (Anti-RBD) and anti-nucleocapsid SARS-CoV-2 IgG antibodies were measured using an automated enzyme-linked immunosorbent assay (ELISA) as previously described (35, 36). Antibody levels were reported as relative ratios to a synthetic standard included as a calibration curve on each assay plate. VHH72-hFc1X7 (VHH-72-Fc) was used as the synthetic standard for anti-spike and anti-RBD antibodies as described previously (35), while human anti-nucleocapsid IgG (clone HC2003, GenScript, no. A02039) was used for anti-nucleocapsid antibody measurements. For VHH-72-hFc1X7, the llama single-domain monoclonal antibody VHH-72 was expressed as a human Fc fusion: VHH-72-hFc1X7 (PDB entry 6WAQ_1). Additional VHHs (NRCoV2–04 and NRCoV2–20) were isolated in-house from llamas immunized with recombinant SARS-CoV-2 trimeric spike ectodomain SmT1. VHH sequences were fused to an antibody-dependent cell-mediated cytotoxicity attenuated human IgG1 Fc domain (hFc1X7, US patent 2019 352 383A1). Our ELISA-based assay was harmonized with the WHO international standard unit – the Binding Antibody Unit (BAU). A table for conversion from relative ratios to BAU/mL is provided in **Supplementary Table 4**.

Thresholds for seropositivity (seroconversion) were determined as ≥3 standard deviations (SDs) from the log means of aggregated data from 300 pre-SARS-CoV-2 negative controls (collected prior to November 2019) (35). Seroconversion / seropositivity thresholds were 0.19, 0.186 and 0.396 for anti-spike, anti-RBD and anti-nucleocapsid antibodies, respectively. We considered the median levels of convalescent serum (taken 21–115 days after symptom onset in a cohort of 211 patients in the general population with a median age 59 years who had mild to severe SARS-CoV-2), as reflective of a robust antibody response (35, 36); the medians were 1.38, 1.25 and 1.13 for anti-spike, anti-RBD, and anti-nucleocapsid antibodies, respectively.

### Spike-pseudotyped lentivirus neutralization assay

Spike cDNAs encoding full-length wild-type SARS-CoV-2 bearing the D614G mutation, and full-length Beta (B.1.351) and Delta (B.1.617.2) variants were obtained from Twist Bioscience (San Francisco, CA). Spike cDNA encoding the full-length Omicron (B.1.1.529 (BA.1)) variant was obtained from Thermo Fisher Scientific (Waltham, MA). The spike expression constructs and pseudotyped lentivirus particles were generated in-house as described previously (30) and are freely available through CoVaRR-Net (https://nbcc.lunenfeld.ca/resources/). The neutralization assay was adapted from a previously validated SARS-CoV-2 spike pseudotyped lentivirus assay with constructs for Beta, Delta and Omicron variants, with minor modifications (59). Briefly, the viral packaging (psPAX2, Addgene, Waterntown, MA), the ZsGreen and luciferase reporter (pHAGE-CMV-Luc2-IRES-ZsGreen-W, kindly provided by Jesse Bloom) and the spike protein constructs were co-transfected into HEK293TN cells using jetPRIME transfection reagent (Polyplus, Illkirch, France). Pseudovirus particles were harvested 48 hours post transfection, and filtered (0.45 µm filters) prior to aliquoting and storage at -80°C. The titre of each pseudovirus was determined by infecting HEK293T-ACE2/TMPRSS2 cells, and a virus dilution resulting in ∼15% infection and > 1,000 relative luciferase units over the control was used in the neutralization assay (1:15 – 1:165 dilution of virus stock, ∼400K relative luciferase units per pseudovirus).

The neutralization assay was conducted by first heat-inactivating (56°C, 30 mins) patient sera samples, then serially diluting sera in assay media at 3-fold (2.5-fold for Omicron testing) over 7 dilutions, starting at 1:22.5 dilution (1:20 for Omicron testing), and incubated with diluted virus at a 1:1 ratio for 1 hour at 37°C prior to addition to HEK293T-ACE2/TMPRSS2 cells. Following the 48 hours incubation, the infected cells were lysed using the BrightGlo Luciferase Assay System (Promega, Madison, WI) and the luminescence signals were quantified using a PerkinElmer Envision instrument. The 50% neutralization titres (ID50s) were calculated in GraphPad Prism 9 (GraphPad Software, San Diego, CA, USA) using a nonlinear regression (log[inhibitor] versus normalized response – variable slope) algorithm. Both the HEK293TN and HEK293T-ACE2/TMPRSS2 cells were maintained at 85% confluency for no more than 25 passages. The dynamic range of the assay is 1:45-1:98,415 (1:40-1:24,414 for Omicron B.1.1.529 (BA.1)) for the detection of neutralizing capabilities of patient sera. Samples not meeting the minimum threshold were considered to be non-neutralizing.

### Statistical analysis

Demographic and clinical characteristics of participants were compared using Wilcoxon rank sum test for continuous variables and Fisher’s exact test for categorical variables.

For paired samples, differences in anti-RBD and anti-spike antibody levels were compared using the Wilcoxon signed-ranks test, differences in proportions (e.g. response categories, proportion with neutralizing antibodies) were assessed using McNemar’s test with continuity correction. ROC analysis was performed using the pROC package, and the optimal decision threshold for each analysis was identified using the “closest.topleft” metric (60).

Antibody titre calculations were performed in GraphPad Prism 9 (GraphPad Software, San Diego, CA, USA), all subsequent analyses were performed in R, version 4.1.2 (61).

### Study approval

This study was registered with Clinical Trials Ontario and was approved by both the Sunnybrook Health Sciences Centre Research Ethics Board and locally by the St. Michael’s Hospital Research Ethics Board with the Project ID 3604. All participants provided written informed consent prior to study participation.

## Data Availability

All or portions of the de-identified data are available upon reasonable request to the authors.

## Author Contributions

DAY, MAH, ACG, KY and CMM participated in study design. DAY, MJO, AL and MAH obtained funding. DAY, MAH and ACG provided supervision. QH and KTA performed the antibody quantification assays and related data acquisition and data analysis. CMM collated clinical data, analyzed the data, made figures and wrote the manuscript. All authors contributed to data interpretation, provided critical input to the manuscript, and approved the final version.

## Data Availability

All or portions of the de-identified data are available upon reasonable request to the corresponding author, Dr. Caitríona McEvoy, or the senior author, Dr. Darren Yuen.

## Acknowledgements

CMM is supported by the Dr. Mary Papantony Nephrology research fund. KTA is supported by a Frederick Banting and Charles Best Canada Graduate Scholarship Doctoral Award. ACG is pillar lead for CoVaRR-Net and the Canada Research Chair (Tier 1) in Functional Proteomics. DAY was supported by a CIHR New Investigator Award. This work was supported by funds from the COVID Immunity Task Force (CITF) (grant No. 2122-HQ-000071) which is funded by the Government of Canada, and the St. Michael’s Hospital Foundation. The robotics equipment used for the ELISA assays is housed in the Network Biology Collaborative Centre (NBCC) at the Lunenfeld-Tanenbaum Research Institute (ACG), a facility supported by Canada Foundation for Innovation funding, by the Ontario Government and by Genome Canada and Ontario Genomics (OGI-139). Sample intake and automated ELISA were performed by Geneviève Mailhot and Melanie Delgado Brand, and members of the NBCC, supervised by Karen Colwill, facilitated sample acquisition and data processing. We would like to thank the Nephrology Clinical Research team at St. Michael’s Hospital, Unity Health Toronto (Michelle Nash, Niki Dacouris, Lindita Dapi, Weiqiu Yuan, and Tiffany Thai) for collection and storage of study samples, and maintenance of the clinical research database. We would also like to thank the St. Michael’s Hospital Kidney Transplant Clinic staff for their support, and finally all of the kidney transplant recipients for their participation in this study.

## Notes

### Competing Interest Statement

All authors have completed the ICMJE uniform disclosure form at www.icmje.org/coi_disclosure.pdf.
ACG has received research funds from a research contract with Providence Therapeutics Holdings, Inc for other projects not directly related to this manuscript. KY has received honoraria from Astra Zeneca. AL has received research funds/contracts from AstraZeneca; Certa; Otsuka; Reata; Retrophin; The George Institute; Chinook Therapeutics; Boeingher Ingelheim; CIHR; Janssen; Merck; Oxford Clinical Trials; Kidney Foundation of Canada; Merck; Amgen; Bayer; Johnson and Johnson; Retrophin; Reata; National Institute of Health; NIDDK. AL has received honoraria from GSK, Takeda; Reata; Astra Zeneca and Janssen. MJO has received honoraria from Baxter Healthcare, and serves on advisory boards for Janssen and Amgen.
No other authors have conflicts of interest to disclose.

### Funding Statement

This work was funded by the COVID Immunity Task Force which is supported by the Government of Canada, and the St. Michael's Hospital Foundation.

### Author Declarations

Ethics committee of St Michaels Hospital gave ethical approval for this work

