## Supplemental Material for "Humoral Responses in the Omicron Era following Three-Dose SARS-CoV-2 Vaccine Series in Kidney Transplant Recipients"

### Table of Contents:

#### Supplemental Tables:

**Supplemental Table 1**: Characteristics of the Healthy Controls
**Supplemental Table 2**: Summary of binding and neutralizing antibody profiles of healthy controls prior to, and at 1 Month following third mRNA SARS-CoV-2 vaccine dose.
**Supplemental Table 3**: Receiver operating characteristic curve and optimal cut-off analysis
**Supplemental Table 4**: Conversion of relative ratios to international units (BAU/mL) for plasma or serum.

#### Supplemental Figures:

**Supplemental Figure 1**: Nucleocapsid Ab profile, and correlation between anti-spike and anti-RBD at Month 1
**Supplemental Figure 2**: Anti-RBD and Anti-spike antibody levels 1 month after mRNA-1273 (Moderna), BNT162b2 (Pfizer- BioNTech), or mixed vaccination.
**Supplemental Figure 3**: Relationship between Month 1 binding antibody response and antibody decline at Month 3
**Supplemental Figure 4**: Correlation between anti-spike and anti-RBD and Nucleocapsid Ab profile at Month 3
**Supplemental Figure 5**: Detection of neutralization antibodies against SARS-CoV-2 wild-type, Beta, Delta and Omicron (BA.1) variants 1 month after mRNA-1273 (Moderna), BNT162b2 (Pfizer-BioNTech), or mixed vaccination.
**Supplemental Figure 6**: Fold-reduction in neutralizing antibodies for all variants compare to wild-type.
**Supplemental Figure 7**: Comparison of binding antibody levels detected in healthy controls and kidney transplant recipients pre and post-third vaccine dose.
**Supplemental Figure 8**: Fold-reduction in neutralizing antibodies for all variants compared to wild-type in healthy controls.
**Supplemental Figure 9**: Differences in anti-RBD and anti-spike antibody levels between responders and non- responders.
**Supplemental Figure 10**: Correlations between anti-spike/RBD IgG responses and neutralizing antibodies.

**Supplemental Table 1: Characteristics of the Healthy Controls (n=13)**

| | | |
|---|---|---|
| Male sex | n (%) | 4 (30.7) |
| Age (years) | Median [IQR] | 46 [31-55] |
| Initial (2-dose) Vaccine Series n (%) | | |
| | Pfizer-BioNTech (BNT162b2) | 13 (100) |
| Dose 3 Vaccine n (%) | | |
| | Pfizer-BioNTech (BNT162b2) | 13 (100) |

Abbreviations: IQR: Inter-quartile range.

**Supplemental Table 2. Summary of binding and neutralizing antibody profiles of healthy controls prior to, and at 1 Month following third mRNA SARS-CoV-2 vaccine dose.**

| **Binding Antibody** | Relative ratio (median [IQR]) | | No. (%) participants with seropositivity | | No. (%) participants with antibody levels ≥ median convalescent level | |
|---|---|---|---|---|---|---|
| | Pre-third dose N=13 | 1 month post 3rd dose N=13 | Pre-third dose N=13 | 1 month post 3rd dose N=13 | Pre-third dose N=13 | 1 month post 3rd dose N=13 |
| Anti-spike | 1.285 [0.860 – 1.378] | 1.825 [1.824 – 1.890] | 13 (100) | 13 (100) | 3 (23.1) | 13 (100) |
| Anti-RBD | 0.669 [0.387 – 0.858] | 1.813 [1.892 – 1.94] | 12 (92.3) | 13 (100) | 1 (7.1) | 13 (100) |
| Anti-nucleocapsid | 0.05 [0.036 – 0.070] | 0.061 [0.055 – 0.078] | 0 (0) | 0 (0) | 0 (0) | 0 (0) |

| **Neutralizing antibody** | $Log_{10}ID_{50}$ (median [IQR]) (of responding patients) | | No. (%) participants with detectable neutralizing antibody | |
|---|---|---|---|---|
| | Pre-third dose N=13 | 1 month post 3rd dose N=13 | Pre-third dose N=13 | 1 month post 3rd dose N=13 |
| Wild Type | 2.163 [1.750 – 2.261] | 3.782 [3.614 – 3.998] | 13 (100) | 13 (100) |
| Beta | 1.737 [1.131 – 2.075] | 3.399 [3.318 – 3.641] | 12 (92.3) | 13 (100) |
| Delta | 1.887 [1.646 – 2.150] | 3.602 [3.282 – 3.720] | 12 (92.3) | 13 (100) |
| Omicron | 1.563 [1.646 – 2.150] | 2.595 [2.538 – 2.646] | 5 (38.5) | 13 (100) |

### Supplemental Table 3: Receiver operating characteristic curve and optimal cut-off analysis

| Variant | Antibody | AUC | 95% CI | Threshold | Specificity | Sensitivity | TN | TP | FN | FP |
|---|---|---|---|---|---|---|---|---|---|---|
| Wild Type | Anti-RBD | 0.99 | 96.74-100% | 0.317 | 100 | 98.88 | 12 | 31 | 1 | 0 |
| | Anti-SmT1 | 0.965 | 89.56-100% | 1.319 | 91.66 | 87.5 | 11 | 28 | 4 | 1 |
| Beta (B.1.351) | Anti-RBD | 0.998 | 99.21-100% | 0.932 | 100 | 96 | 19 | 24 | 1 | 0 |
| | Anti-SmT1 | 0.987 | 96.56-100% | 1.481 | 94.74 | 96 | 18 | 24 | 1 | 1 |
| Delta (B.1.617.2) | Anti-RBD | 1 | NA | 0.627 | 100 | 100 | 16 | 28 | 0 | 0 |
| | Anti-SmT1 | 0.978 | 94.39-100% | 1.391 | 93.75 | 89.29 | 15 | 25 | 3 | 1 |
| Omicron (B.1.1.529) | Anti-RBD | 0.91 | 80.82-100% | 1.198 | 87.5 | 95 | 21 | 19 | 1 | 3 |
| | Anti-SmT1 | 0.925 | 85.22-99.78% | 1.475 | 83.33 | 85 | 20 | 17 | 3 | 4 |

Abbreviations: AUC: area under the curve; CI: confidence intervals; FN: false negative; FP: false positive; NA: not applicable; RBD: receptor binding domain; Anti-SmT1: Anti-spike; TN: true negative; TP: true positive.

**Supplemental Table 4. Conversion of relative ratios to international units (BAU/mL) for plasma or serum.**

| Nucleocapsid | | | | | | |
|---|---|---|---|---|---|---|
| Dilution fold (d) | | 160 | | 640 | | Note |
| Volume (ul sample/well) | | 0.0625 | | 0.015625 | | |
| Relative Ratio (RR) | $log_2$(RR) | $log_2$(BAU/mL) | BAU/ml | $log_2$(BAU/mL) | BAU/ml | |
| 2 | 1.0 | 8.38 | 333.98 | 10.38 | 1,335.94 | Upper limit of linear range |
| 1 | 0.0 | 6.98 | 126.34 | 8.98 | 505.34 | |
| 0.5 | -1.0 | 5.58 | 47.79 | 7.58 | 191.15 | |
| 0.396 | -1.3 | 5.11 | 34.46 | 7.11 | 137.83 | Positivity threshold |
| 0.25 | -2.0 | 4.18 | 18.08 | 6.18 | 72.31 | |
| 0.125 | -3.0 | 2.77 | 6.84 | 4.77 | 27.35 | |
| 0.0625 | -4.0 | 1.37 | 2.59 | 3.37 | 10.35 | Lower limit of linear range |
| Formula: log2(BAU mL-1 @ sample dilution fold d) = (log2(RR) - 0.243)/ 0.713 + log2(d) | | | | | | |

| RBD | | | | | | |
|---|---|---|---|---|---|---|
| Dilution fold (d) | | 160 | | 640 | | Note |
| Volume (ul sample/well) | | 0.0625 | | 0.015625 | | |
| Relative Ratio (RR) | $log_2$(RR) | $log_2$(BAU/mL) | BAU/ml | $log_2$(BAU/mL) | BAU/ml | |
| 1 | 0.0 | 8.12 | 278.37 | 10.12 | 1,113.50 | Upper limit of linear range |
| 0.5 | -1.0 | 6.82 | 112.63 | 8.82 | 450.51 | |
| 0.25 | -2.0 | 5.51 | 45.57 | 7.51 | 182.27 | |
| 0.186 | -2.4 | 4.95 | 30.97 | 6.95 | 123.89 | Positivity threshold |
| 0.125 | -3.0 | 4.20 | 18.44 | 6.20 | 73.74 | |
| 0.0625 | -4.0 | 2.90 | 7.46 | 4.90 | 29.84 | |
| 0.03125 | -5.0 | 1.59 | 3.02 | 3.59 | 12.07 | Lower limit of linear range |
| Formula: log2(BAU mL-1 @ sample dilution fold d) = (log2(RR) + 0.612)/ 0.766 + log2(d) | | | | | | |

| Spike | | | | | | |
|---|---|---|---|---|---|---|
| Dilution fold (d) | | 160 | | 640 | | Note |
| Volume (ul sample/well) | | 0.0625 | | 0.015625 | | |
| Relative Ratio (RR) | $log_2$(RR) | $log_2$(BAU/mL) | BAU/ml | $log_2$(BAU/mL) | BAU/ml | |
| 1 | 0.0 | 2.55 | 5.86 | 6.55 | 93.80 | Upper limit of linear range |
| 0.5 | -1.0 | 1.28 | 2.42 | 5.28 | 38.75 | |
| 0.25 | -2.0 | 0.00 | 1.00 | 4.00 | 16.01 | |
| 0.19 | -2.4 | -0.50 | 0.70 | 3.50 | 11.28 | Positivity threshold |
| 0.125 | -3.0 | -1.28 | 0.41 | 2.72 | 6.61 | |
| 0.0625 | -4.0 | -2.55 | 0.17 | 1.45 | 2.73 | |
| 0.03125 | -5.0 | -3.83 | 0.07 | 0.17 | 1.13 | Lower limit of linear range |
| Formula: log2(BAU mL-1 @ sample dilution fold d) = (log2(RR) – 0.604)/0.784 + log2(d) | | | | | | |

Abbreviations: BAU: Binding antibody unit. RBD: Receptor binding domain.

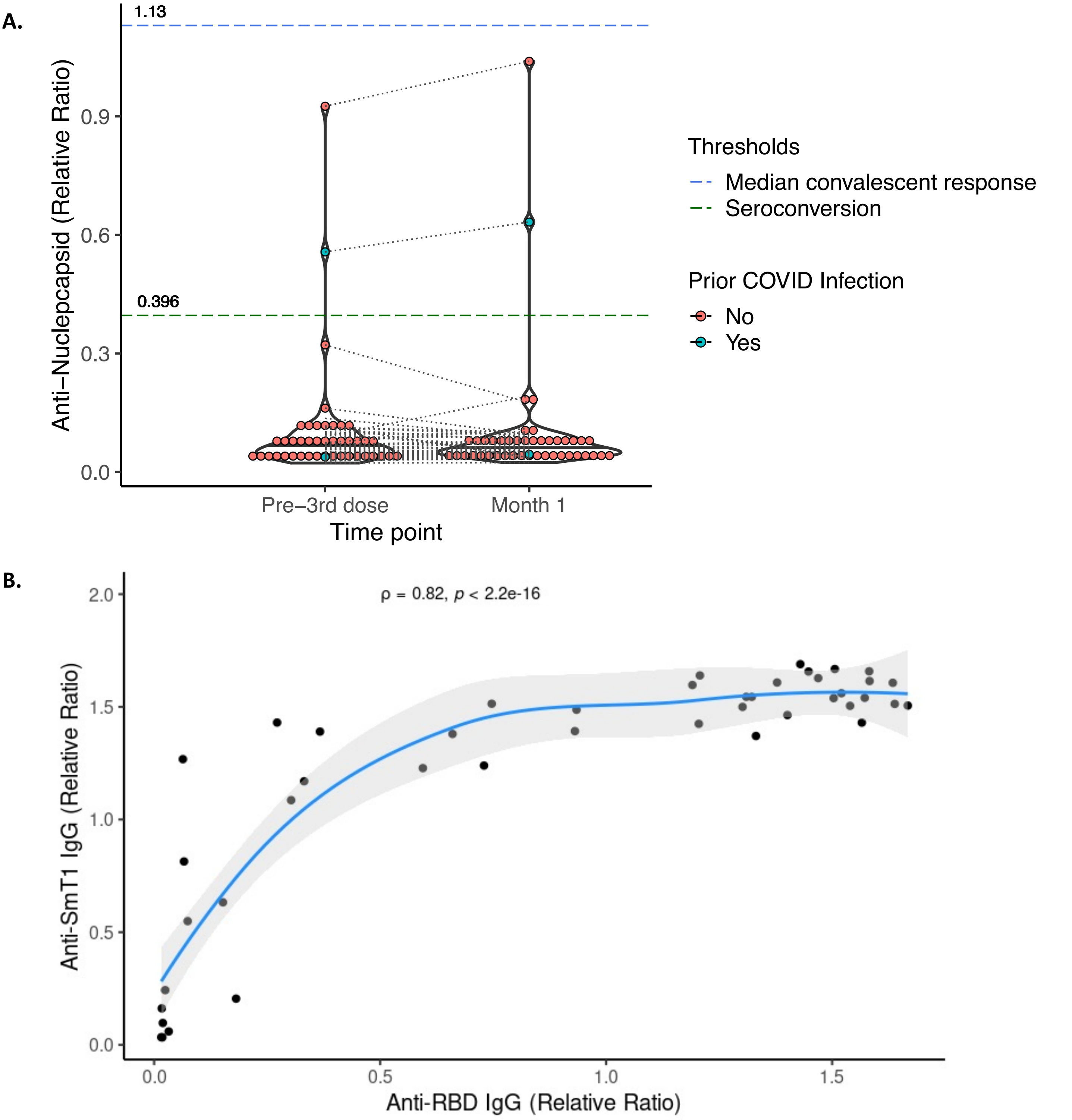


**Supplemental Figure 1: Nucleocapsid Ab profile, and correlation between anti-spike and anti-RBD at Month 1**
**A)** Levels of serum anti-nucleocapsid IgG in the 44 participants with bloods drawn prior to the third dose, and at 1 month post-third-dose. The values depicted are relative rations against a synthetic standard. Threshold lines and values demonstrate seropositivity (green dashed line) and the median convalescent response (blue dashed line). Individual values are colored to depict prior COVID-19 infection as shown in the legend. **B)** Correlation between serum anti-RBD and anti-spike antibody levels at 1 month following third mRNA vaccine dose. Anti-RBD and anti-spike antibodies are expressed as relative ratios. The blue line indicates the regression line fit by LOESS, with 95% confidence intervals shaded in light grey. Spearman's rho coefficient and associated P value are marked.

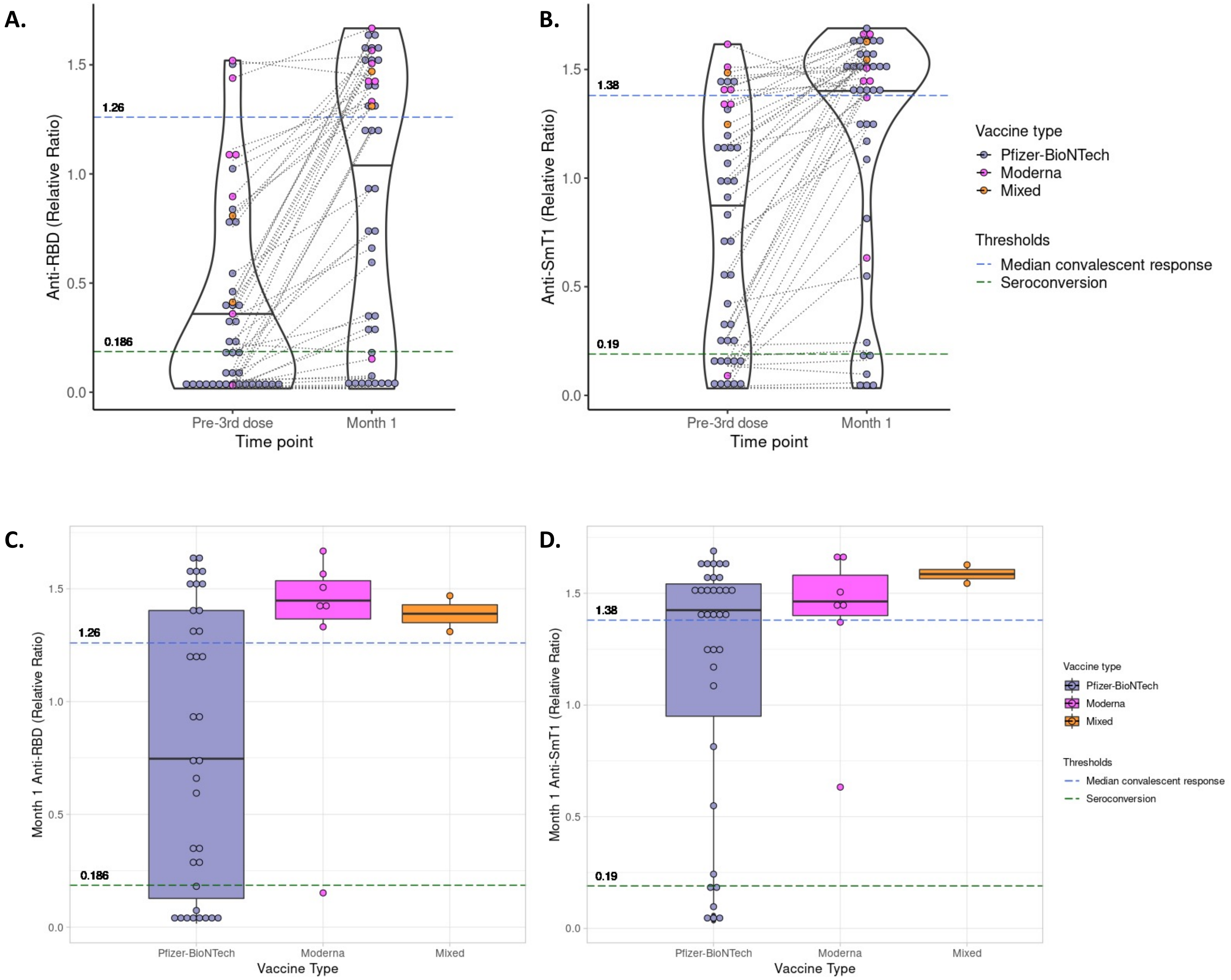


**Supplemental Figure 2: Anti-RBD and Anti-spike antibody levels 1 month after mRNA-1273 (Moderna), BNT162b2 (Pfizer-BioNTech), or mixed vaccination. A)** Anti-RBD and **B)** Anti-spike antibodies, detected in 44 participants with blood samples drawn pre- and at 1 month post-third dose. The vaccine type administered is denoted by the coloured dots, as outlined in the figure legend. The impact of vaccine type on the likelihood of having a robust response at Month 1 was assessed using Fisher's exact test with post-hoc pairwise comparisons as indicated. RBD p= 0.001 (post-hoc pairwise comparisons: Pfizer-BioNTech - Moderna adjusted p value =0.09, Pfizer-BioNTech - Mixed vaccine, adjusted p value = 0.14, Moderna - Mixed Vaccine adjusted p value =1); Anti-spike p= 0.59. **C)** Anti-RBD and **D)** anti-spike antibody levels are plotted between groups of patients who received Moderna, Pfizer-BioNTech, or mixed vaccination, as indicated. Significance was assessed using Kruskal-Willis test, p= 0.1 (anti-RBD) and p=0.22 (anti-spike) .
For all images: The values depicted are relative ratios against a synthetic standard. Serum volume 0.0625 µL. Threshold lines and marked values demonstrate seropositivity (green dashed line) and the median convalescent response (blue dashed line). Solid black lines indicate the median ratio values for each grouping.

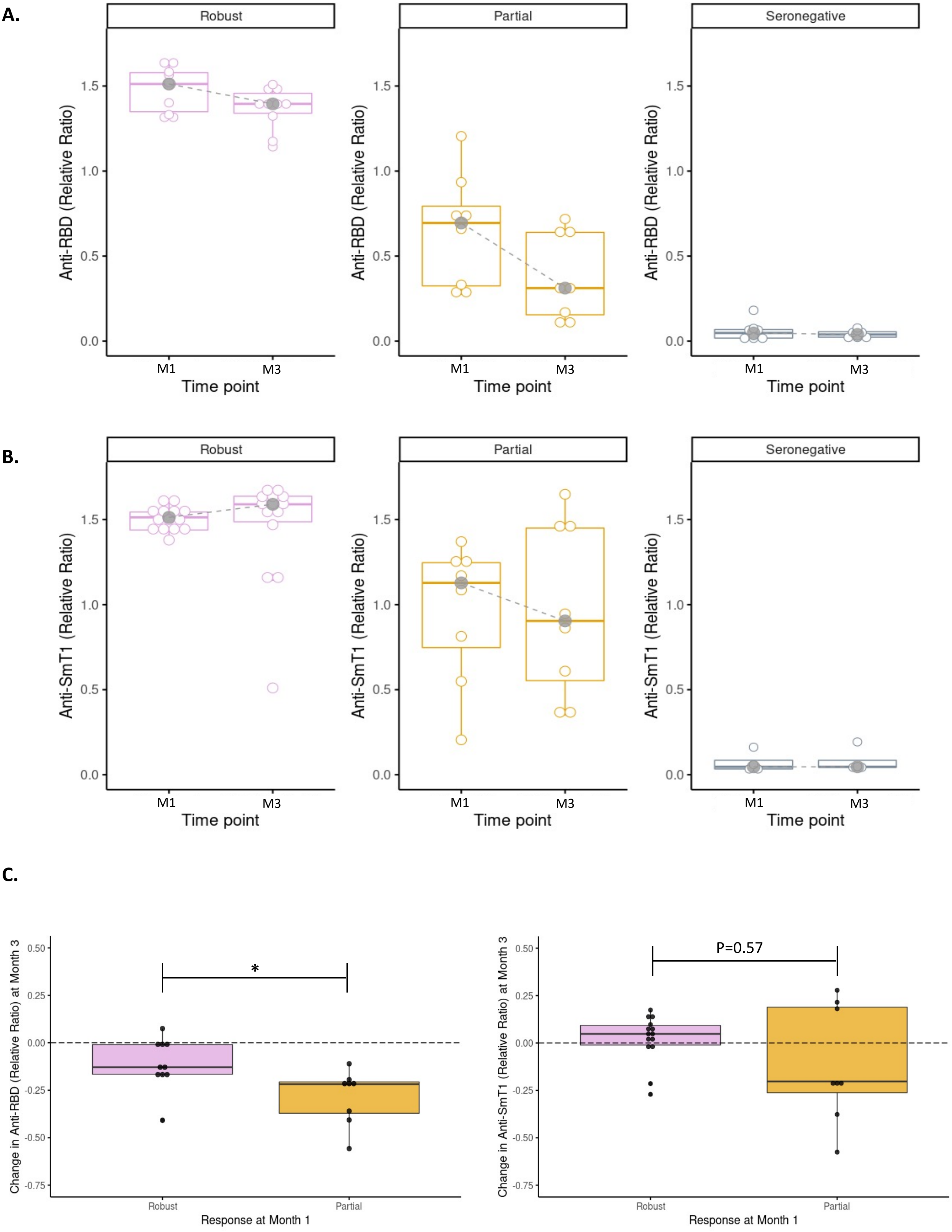


**Supplemental Figure 3**: Month 1 and Month 3 antibody levels of **A)** anti-spike and **B)** anti-RBD antibodies in the 44 participants with blood samples drawn pre- and at 1 month post-third dose, categorized based on the participant's response category as assessed on Month 1 bloods: Robust (above the threshold of the median convalescent response in healthy controls); Partial (above the threshold for seropositivity, but below the threshold of robust response), and Seronegative (below the threshold for seropositivity). **C)** Absolute change in anti-RBD (left) and anti-spike (right) antibody levels between Month 1 and Month 3. Patients are grouped according to their response category at Month 1 (Robust or Partial). Solid black lines indicate the median ratio values for each grouping. Significance assessed using Wilcoxon rank sum test. * $P \leq 0.05$, **$P \leq 0.01$, ***$P \leq 0.001$.

A.

B.

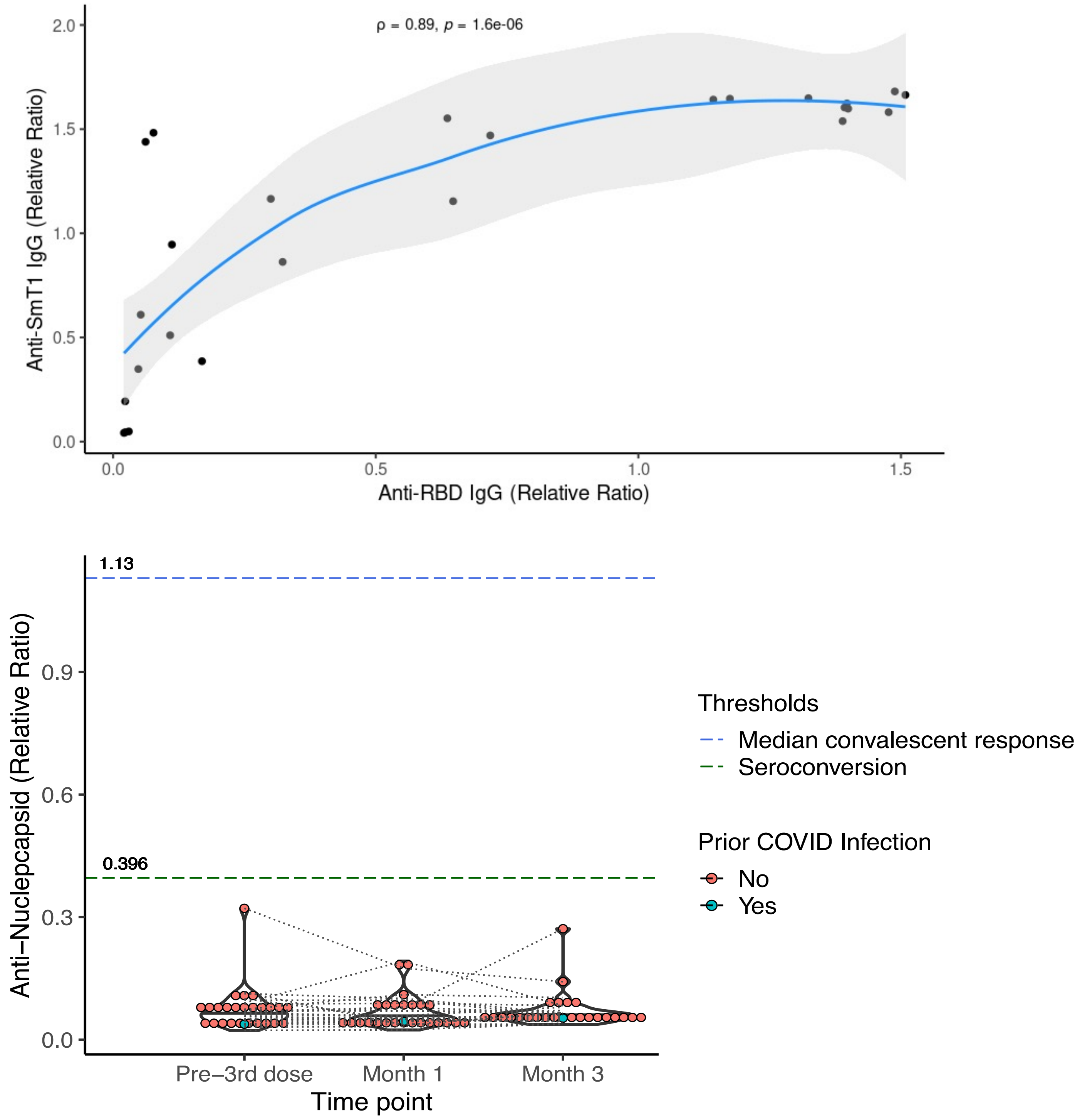


**Supplemental Figure 4: Correlation between anti-spike and anti-RBD and Nucleocapsid Ab profile at Month 3**
**A)** Correlation between serum anti-RBD and anti-spike antibody levels at 3 months following third mRNA vaccine dose. Anti-RBD and anti-spike antibodies are expressed as relative ratios. The blue line indicates the regression line fit by LOESS (locally estimated scatterplot smoothing), with 95% confidence intervals shaded in light grey. Spearman's rho coefficient and associated P value are marked. **B)** Levels of serum anti-nucleocapsid IgG in the 26 participants with blood samples drawn prior to the third dose, and at 1 and 3 months post-third-dose. The values depicted are relative rations against a synthetic standard. Threshold lines and values demonstrate seropositivity (green dashed line) and the median convalescent response (blue dashed line). Individual values are colored to depict prior COVID-19 infection as shown in the legend.

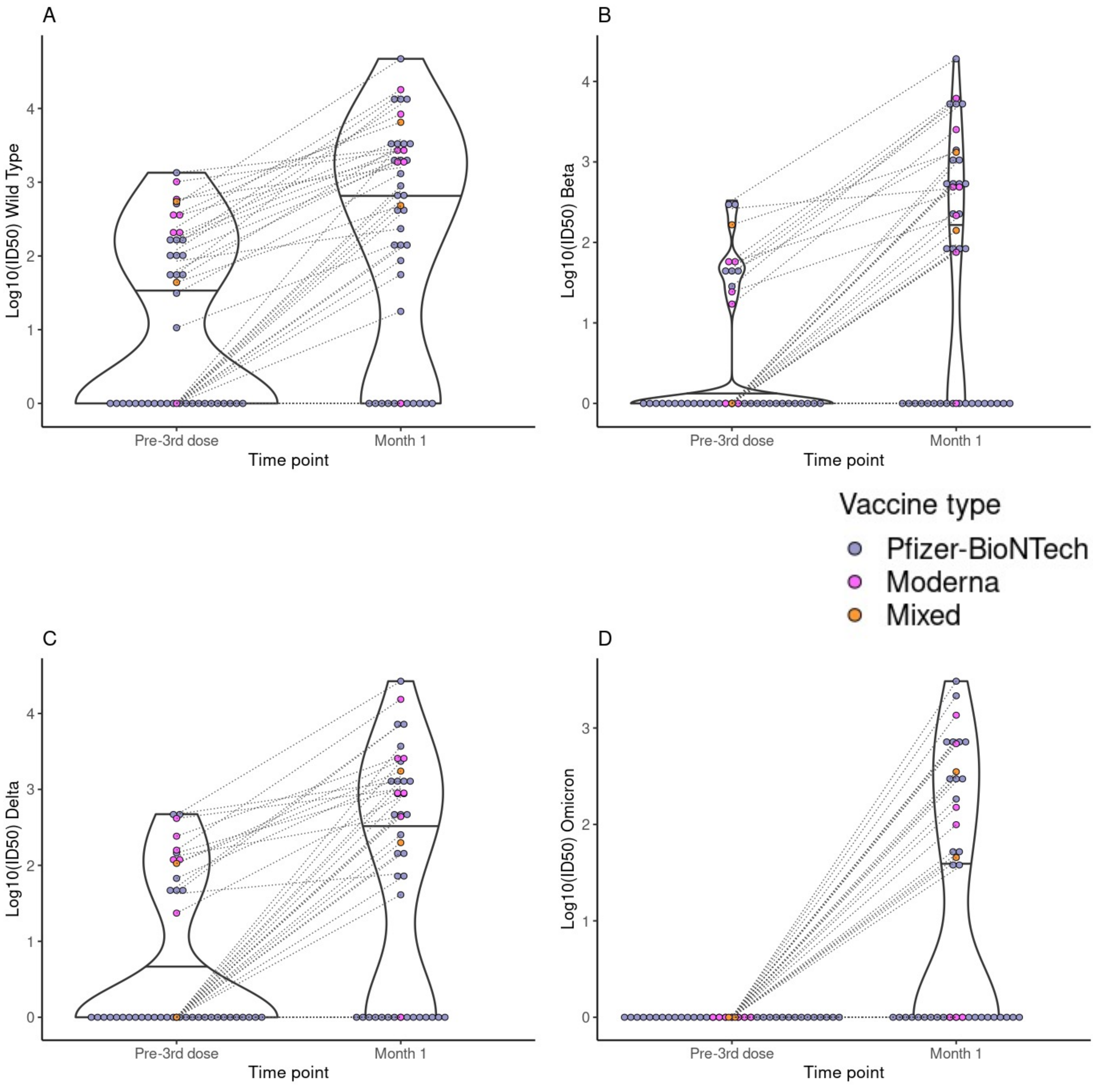


**Supplemental Figure 5: Detection of neutralization antibodies against SARS-CoV-2 wild-type, Beta, Delta and Omicron (BA.1) variants 1 month after mRNA-1273 (Moderna), BNT162b2 (Pfizer-BioNTech), or mixed vaccination.** Neutralizing antibodies ($Log_{10}ID_{50}$)against **A)** wild type (WT), **B)** Beta, **C)** Delta and **D)** Omicron variants, detected in 44 participants with blood samples drawn pre- and at 1 month post-third dose. The impact of vaccine type on the numbers of responders ($Log_{10}ID_{50}$ >0) and non-responders (undetectable neutralizing antibody) at Month 1 was assessed using Fisher's exact test, p = 0.81 (WT), p = 0.15 (Beta), p= 0.252 (Delta) and p= 0.208 (Omicron). For all images: Vaccine type is indicated in the figure legend. Paired values are linked with black dashed lines. Solid black lines in each violin plot indicate the median $Log_{10}ID_{50}$ values for each variant.

**A.**

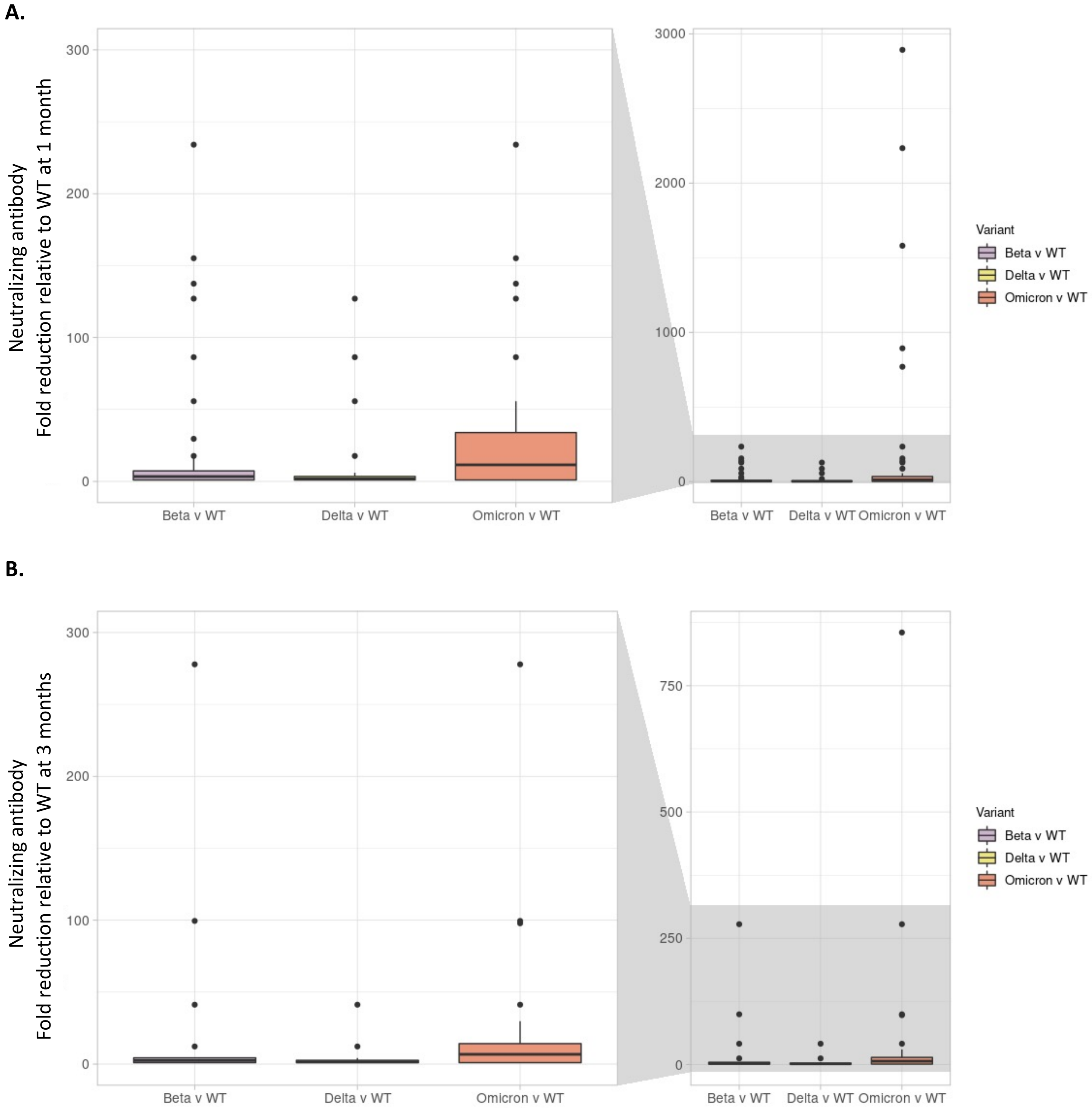


**Supplemental Figure 6: Fold-reduction in neutralizing antibodies for all variants compare to wild-type.** Fold reduction in ID50 for Beta, Delta and Omicron compared to wild-type (WT) virus. Fold reduction for each participant was calculated by dividing the absolute ID50 value for the WT virus by the absolute ID50 value for the variant. The height of the bar represents the median value and error bars represent the interquartile range of values. Fold-reduction is shown for **A)** 1-month post-third dose samples (n = 44) **B)** and 3-month post-third dose samples (n = 26). For A) and B) the graphs on the left are a more expanded view of the shaded portion of the graphs on the right hand side, which depict the entire range of values.

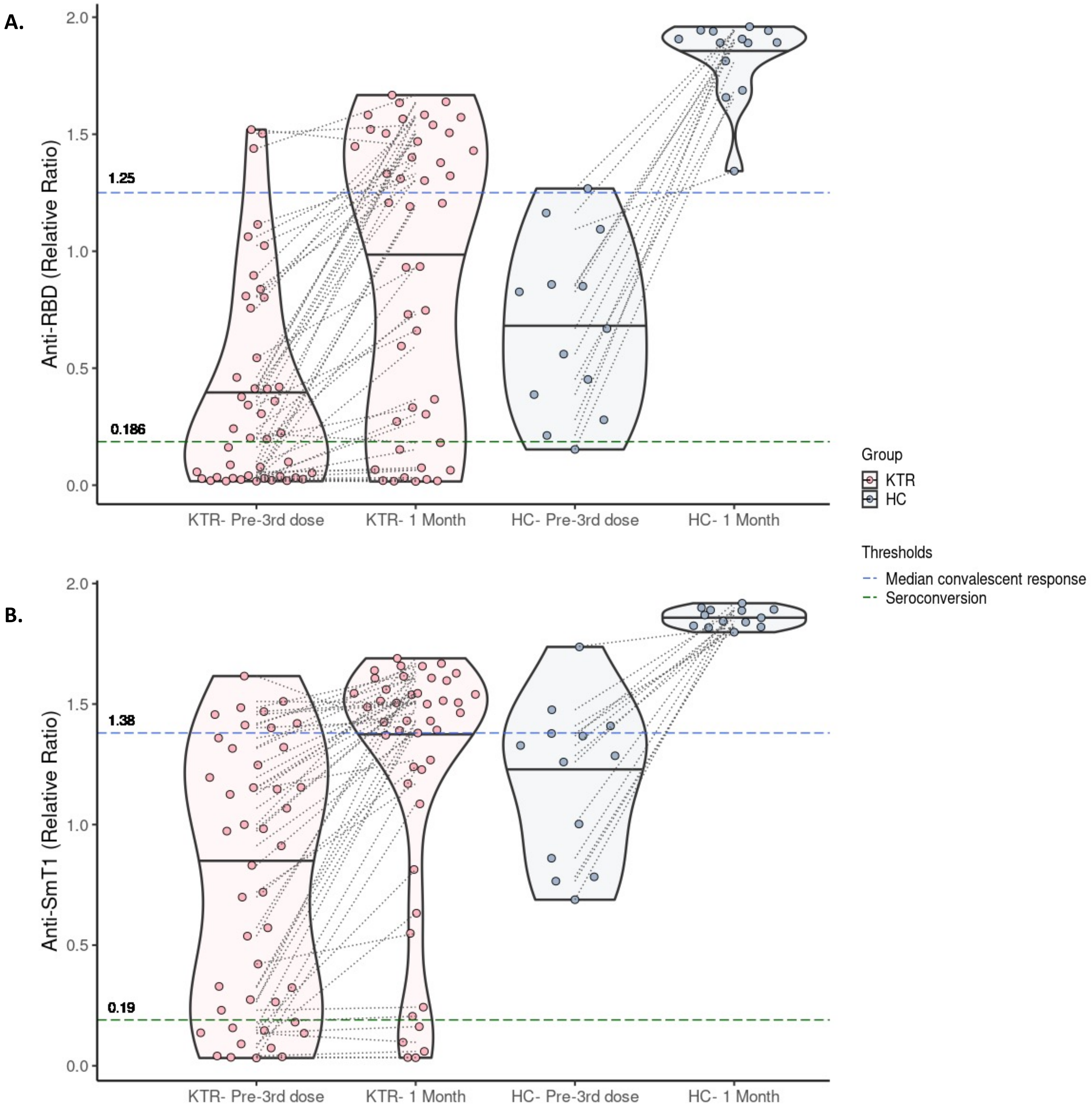


**Supplemental Figure 7: Comparison of binding antibody levels detected in healthy controls and kidney transplant recipients pre and post-third vaccine dose. A)** Anti-spike and **B)** anti-RBD antibodies detected in 44 kidney transplant recipients (KTRs) and 13 healthy controls (HC) before the third vaccine dose and at month 1 post-third dose. For all images: Paired values are linked with black dashed lines. Solid black lines in each violin plot indicate the median values for each antibody.

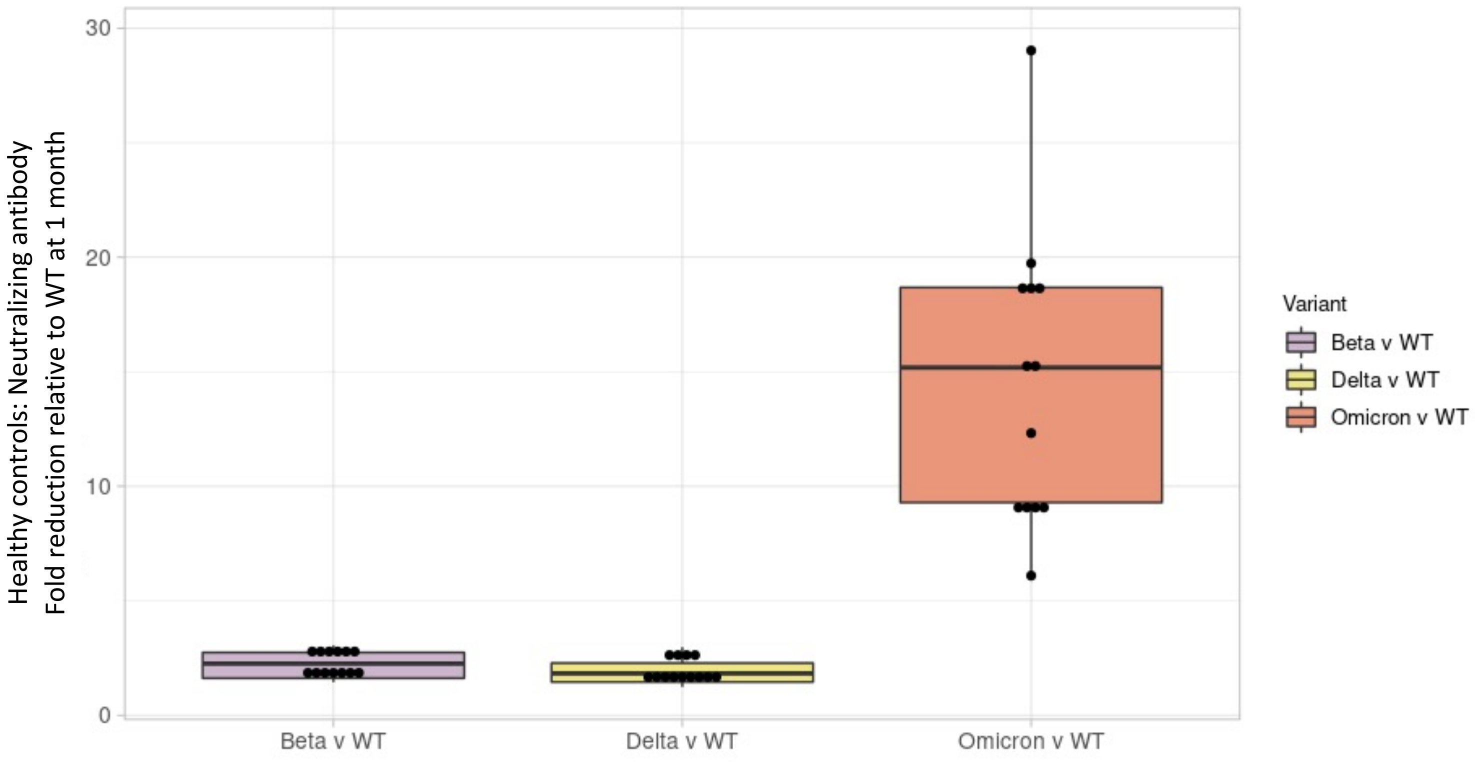


**Supplemental Figure 8: Fold-reduction in neutralizing antibodies for all variants compared to wild-type in healthy controls.**

Fold reduction in ID50 for Beta, Delta and Omicron compared to wild-type (WT) virus. Fold reduction for each participant was calculated by dividing the absolute ID50 value for the WT virus by the absolute ID50 value for the variant. The height of the bar represents the median value and error bars represent the interquartile range of values.

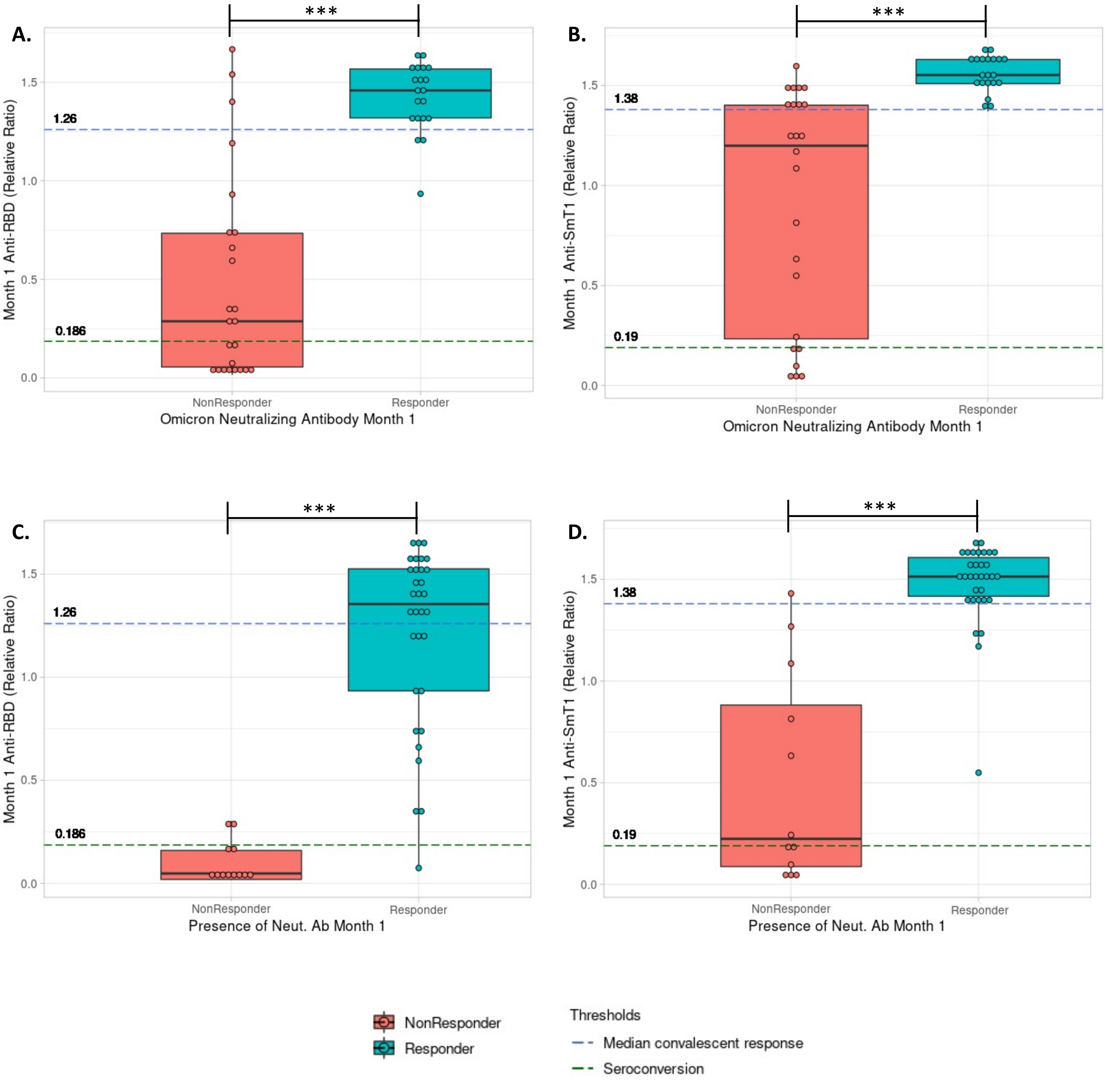


**Supplemental Figure 9: Differences in anti-RBD and anti-spike antibody levels between responders and non-responders.**

**A)** Anti-RBD and **B)** anti-spike antibody levels are plotted between groups of patients who responded (detectable neutralizing antibody against Omicron) or did not respond (zero Omicron-specific neutralizing antibody detected) at one month following third vaccine dose. **C)** Anti-RBD and **D)** Anti-spike antibody levels of patients who had detectable neutralizing antibody against one or more of the variants tested, compared to those who had zero detectable neutralizing antibody against all 4 variants (WT, Beta, Delat and Omicron). Median values are marked with a black line. Differences in means were assessed using Wilcoxon rank sum test. * P ≤ 0.05, **P ≤ 0.01, ***P ≤ 0.001.

A.

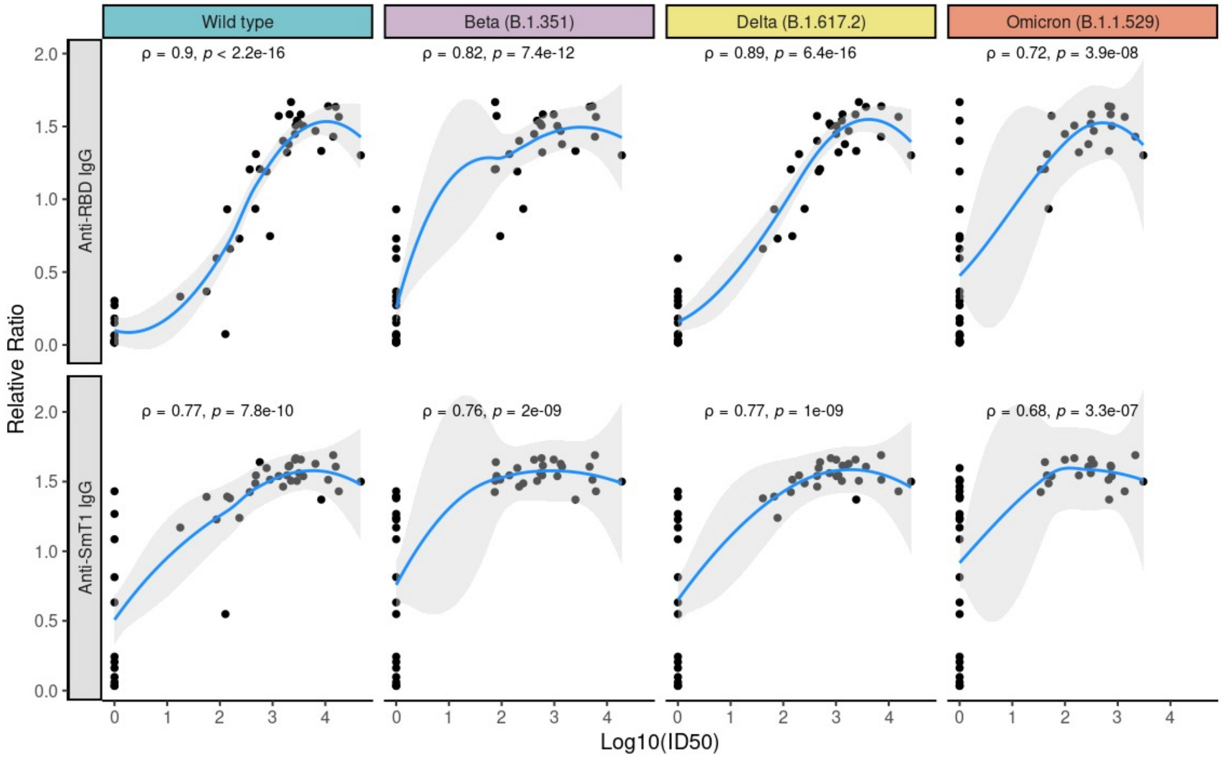


B.

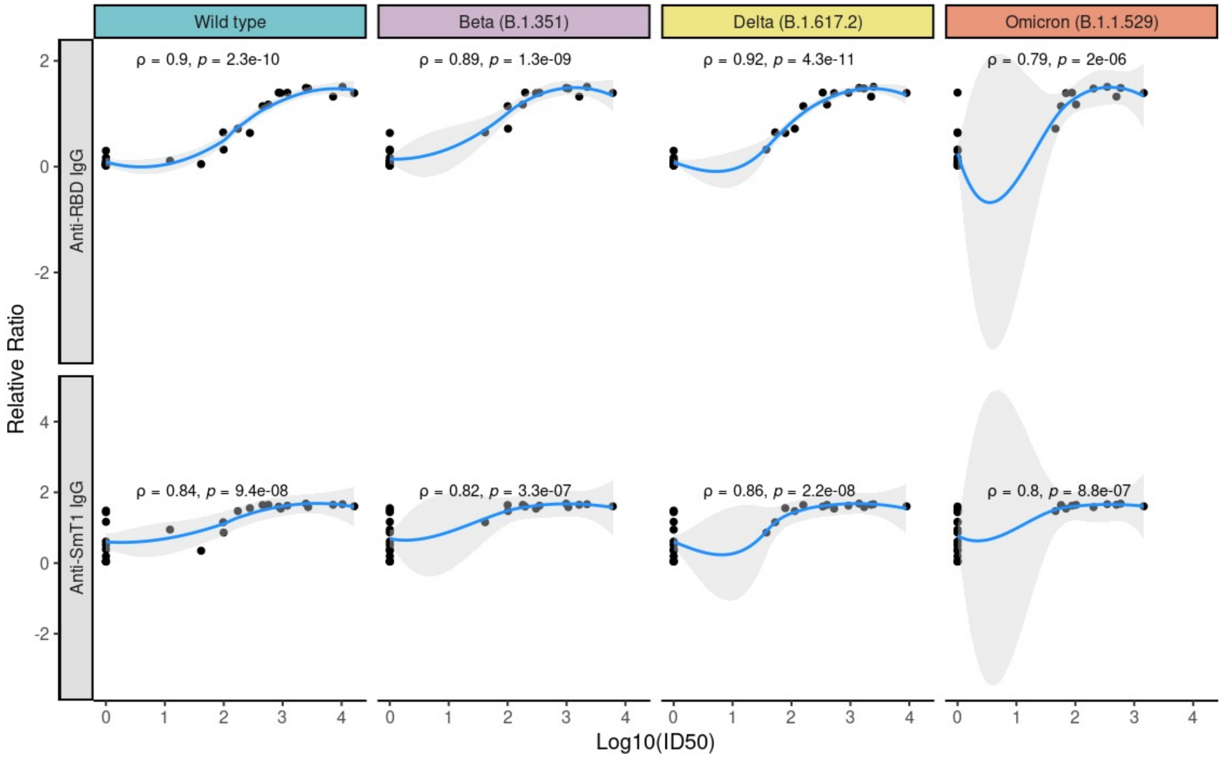


**Supplemental Figure 10: Correlations between anti-spike/RBD IgG responses and neutralizing antibodies**.
A) Correlation between anti-RBD and anti-spike IgG antibody (relative ratio) one month following third vaccine dose and neutralizing antibodies ($log_{10}ID_{50}$) at **A)** 1 month (n=44 patients) and **B)** 3 months (n= 26 patients) following the third dose. The blue line indicates the regression line fit using LOESS (locally estimated scatterplot smoothing), and grey shading indicates the 95% confidence interval. Spearman's rho coefficients and associated P values are indicated.
